# What would other emergency stroke teams do? Using explainable machine learning to understand variation in thrombolysis practice

**DOI:** 10.1101/2023.04.24.23289017

**Authors:** Kerry Pearn, Michael Allen, Anna Laws, Thomas Monks, Richard Everson, Martin James

## Abstract

**Objectives:** To understand between-hospital variation in thrombolysis use among patients in England and Wales who arrive at hospital within 4 hours of stroke onset.

**Design:** Machine learning was applied to the Sentinel Stroke National Audit Programme (SSNAP) data set, to learn which patients in each hospital would likely receive thrombolysis.

**Setting:** All hospitals (n=132) providing emergency stroke care in England and Wales. Thrombolysis use in patients arriving within 4 hours of known or estimated stroke onset ranged from 7% to 49% between hospitals.

**Participants:** 88,928 stroke patients recorded in the national stroke audit who arrived at hospital within 4 hours of stroke onset, from 2016 to 2018.

**Intervention:** Extreme Gradient Boosting (XGBoost) machine learning models, coupled with a SHAP model for explainability.

**Main Outcome Measures:** Shapley (SHAP) values, providing estimates of how patient features, and hospital identity, influence the odds of receiving thrombolysis.

**Results:** The XGBoost/SHAP model revealed that the odds of receiving thrombolysis reduced 9 fold over the first 120 minutes of arrival-to-scan time, varied 30 fold depending on stroke severity, reduced 3 fold with estimated rather than precise stroke onset time, fell 6 fold with increasing pre-stroke disability, fell 4 fold with onset during sleep, fell 5 fold with use of anticoagulants, fell 2 fold between 80 and 110 years of age, reduced 3 fold between 120 and 240 minutes of onset-to-arrival time, and varied 13 fold between hospitals. The hospital attended explained 56% of the variance in between-hospital thrombolysis use, adding in other hospital processes explained 74%, the patient population alone explained 36%, and the combined information from both patient population and hospital processes explained 95% of the variance in between-hospital thrombolysis use. Patient SHAP values expose how suitable a patient is considered for thrombolysis. Hospital SHAP values expose the threshold at which patients are likely to receive thrombolysis.

**Conclusions:** Using explainable machine learning, we have identified that the majority of the between-hospital variation in thrombolysis use in England and Wales, for patients arriving with time to thrombolyse, may be explained by differences in in-hospital processes and differences in attitudes to judging suitability for thrombolysis.

## 1 Introduction

Stroke remains one of the top three global causes of death and disability ^1^. Despite reductions in age-standardised rates of stroke, ageing populations are driving an increase in the absolute number of strokes^1^. Across Europe, in 2017, stroke was found to cost healthcare systems €27 billion, or 1.7% of health expenditure ^2^. Thrombolysis with recombinant tissue plasminogen activator, can significantly reduce disability after ischaemic stroke, so long as it is given in the first few hours after stroke onset ^3^. Despite thrombolysis being of proven benefit in ischaemic stroke, use of thrombolysis varies significantly both between and within European countries ^4^. In England and Wales the national stroke audit reported that in 2021/22, 20 years on from the original European Medicines Agency licencing of alteplase for acute ischaemic stroke, thrombolysis rates for emergency stroke admissions varied from just 1% to 28% between hospitals ^5^, with a median rate of 10.4% and an inter-quartile range of 8%-13%, against a 2019 NHS England long term plan that 20% of patients of emergency stroke admissions should be receiving thrombolysis ^6^.

Studies have shown that reasons for low and varying thrombolysis rates are multi-factorial. Reasons include late presentation ^4^, lack of expertise ^4^ or lack of clear protocols or training ^7^, delayed access to specialists ^8^, and poor triage by ambulance or emergency department staff ^7^. For many factors, the establishment of primary stroke centres has been suggested to improve the emergency care of patients with stroke and reduce barriers to thrombolysis ^7^, with a centralised model of primary stroke centres leading to increased likelihood of thrombolysis ^9;10;11^.

In addition to organisational factors, clinicians can have varying attitudes to which patients are suitable candidates for thrombolysis. In a discrete choice experiment ^12^, 138 clinicians considered hypothetical patient vignettes, and responded as to whether they would give the patients thrombolysis. The authors concluded that there was considerable heterogeneity among respondents in their thrombolysis decision-making. Areas of difference were around whether to give thrombolysis to mild strokes, to older patients beyond 3 hours from stroke onset, and when there was pre-existing disability.

Based on national audit data from three years of emergency stroke admissions, we have previously built models of the emergency stroke pathway using clinical pathway simulation to examine the potential scale of the effect of changing two aspects of the stroke pathway performance (1. the in-hospital process speeds, and 2. the proportion of patients with a determined stroke onset time), and using machine learning to examine the effect of replicating clinical decision-making around thrombolysis from higher thrombolysing hospitals to lower thrombolysing hospitals ^13;14^. The machine learning model learned whether any particular patient would receive thrombolysis in any particular emergency stroke centre. Using these models we found that it would be credible to target an increase in average thrombolysis in England and Wales, from 11% to 18%, but that each hospital should have its own target, reflecting differences in local populations. We found that the largest increase in thrombolysis use would come from replicating thrombolysis decision-making practice from higher to lower thrombolysing hospitals. Two other important factors influencing thrombolysis rates were determination of stroke onset time in some hospitals, and improving the speed of the in-hospital thrombolysis pathway.

In our previous work we established that we could predict the use of thrombolysis in patients arriving within 4 hours of known stroke onset with 84.3% accuracy ^14^. We could then ask the question “What if this patient attended another hospital - would they likely be given thrombolysis?” As this was a *‘black-box’* decision-forest model we could not effectively explain the relationship between patient level data (‘features’) and their chance of receiving thrombolysis, or identify and explain the features which different hospitals would differ on.

In this paper, therefore, we seek to use *explainable machine learning* to understand the relationship between patient and hospital features and the use of thrombolysis across England and Wales, and we seek to understand how hospitals differ in their attitudes to use of thrombolysis, and how much difference in use of thrombolysis may be explained by those differences. We use an *eXtreme Gradient Boosting model* ^15^ (XGBoost) to make predictions and then use an additional *SHapley Additive exPlanations* ^16^ (SHAP) model to explain the contribution of each feature to the model prediction.

## 2 Methods

All modelling and analysis was performed using Python in Jupyter Notebooks ^17^, with general analysis and plotting performed using NumPy ^18^, Pandas ^19^, Scikit-Learn ^20^, and Matplotlib ^21^.

Further details of methods may also be found in the appendix.

All code, with detailed results, used is available online as a Jupyter book at https://samuel-book.github.io/samuel_shap_paper_1/ and available on GitHub^22^.

### 2.1 Data

Data were retrieved for 246,676 emergency stroke admissions to acute stroke teams in England and Wales for the three calendar years 2016 - 2018, obtained from the Sentinel Stroke National Audit Programme^a^ (SSNAP). Data fields were provided for the hyper-acute phase of the stroke pathway, up to and including our target feature: *receive thrombolysis* (full details of the data fields obtained are provided in the appendix). Of these patients, 88,928 arrived within 4 hours of known (precise or estimated) stroke onset, and were used in this modelling study (as they represent the patients that had time left to receive thrombolysis). The data included 132 acute stroke hospitals (these were all units admitting an average of 100 patients per year, and delivering thrombolysis to at least 10 patients over 3 years). For modelling purposes, the categorical feature *Stroke team* in the SSNAP dataset was represented as 132 one-hot encoded features (a separate feature for each hospital). One-hot encoding is a process which converts a categorical feature to a numerical form. There are 60 original features used from the SSNAP dataset (before one-hot encoding categorical features).

SSNAP has near-complete coverage of all acute stroke admissions in the UK (outside Scotland). All hospitals admitting acute stroke participate in the audit, and year-on-year comparison with Hospital Episode Statistics^b^ confirms estimated case ascertainment of 95% of coded cases of acute stroke.

### 2.2 Machine learning models (to predict thrombolysis use)

We used an *eXtreme Gradient Boosting model* ^15^ (‘XGBoost’) to predict the probability of use of thrombolysis for each patient from their other feature values. We chose XGBoost for its efficiency, and because we working with a system with known non-linear relationships and potential feature interactions.

#### 2.2.1 Feature selection

Before applying SHAP, we aimed to enhance the understandability and explainability of our models through restricting the features (patient level data fields) included in the model.

Features were selected one at a time from the 60 original features in the SSNAP dataset by forward-feature selection ^23^ (identifying one feature at a time that led to the greatest improvement in accuracy). Model accuracy was measured by Receiver Operating Characteristic (ROC) Area Under Curve (AUC), using 5-fold cross-validation. We repeated this process to identify the top 25 features, and used these results to identify the number of features to include in our machine learning models (based on the observed diminishing returns).

#### 2.2.2 Model accuracy

Model accuracy, ROC AUC, sensitivity and specificity were measured using stratified 5-fold cross validation. The appendix contains further model accuracy analysis.

#### 2.2.3 The machine learning models

For the different analysis included in this paper, we trained three XGBoost models:

1. *K-fold model* Description: Train/test five models using all 88,928 patients in a 5-fold cross-validation. Purpose: To robustly test the accuracy of the model, and to test reproducibility of SHAP values across the five k-fold models.
2. *All data model* Description: Train a single model using all 88,928 patients. Purpose: Having understood the model accuracy (from the *k-fold model*), we used all the data to create a single model to understand and explain predicitons.
3. *10k holdout model* Description: Select 10k patients (stratified by hospital and thrombolysis use) to keep in a hold-back dataset, and train a single model using the remaining 78,928 patients. Purpose: By passing all of the 10k patients (that were held back from the training process) through the fitted model, whilst setting the hospital attended feature to each hospital in turn, we predicted the thrombolysis rate for each hospital if all hospitals saw the same patients, revealing the variation in thrombolysis that is caused by the hospital, rather than by between-hospital variation in patient populations.

For all models, a single model was fitted for all hospitals, with hospital attended being a feature (represented as a one-hot encoded feature).

### 2.3 SHapley Additive exPlanation (SHAP) values

We sought to make our models explainable using SHAP values (calculated using the SHAP library ^16^). SHAP provides a measure of the contribution of each feature value to the final predicted probability of receiving thrombolysis for that individual. The SHAP values for each feature are comprised of the feature’s main effect (the effect of that feature in isolation) and all of the pairwise interaction effects with each of the other features (a value per feature pairing). SHAP values provide the influence of each feature as the change in log-odds of receiving thrombolysis. SHAP values expressed as log-odds are additive, i.e. the final log-odds of receiving thrombolysis is the sum of the base model prediction (the log-odds of receiving thrombolysis before feature influences are considered), and the SHAP values for each feature (comprised of the feature’s main effect and all of the pairwise interaction effects with each of the other features).

### 2.4 The relationship between feature values and the odds of receiving thrombolysis

For each feature, we examined the relationship between feature values and their corresponding SHAP values (we used values from the *all data* model).

### 2.5 How much of the between-hospital thrombolysis use can be explained by the identity of the hospital attended?

For each hospital we compared the mean SHAP main effect value for the hospital attended (using values from the *all data* model) with the hospitals observed thrombolysis use. Each hospital has their own patient population.

To reveal the variation in thrombolysis rate due to hospital, rather than patient mix, we also compared the mean hospital attended SHAP main effect value for the identical 10k patient cohort attending each hospital, with the hospitals’ predicted thrombolysis use for this 10k patient cohort (we used values from the *10k holdout* model).

By using the SHAP main effect (instead of the full SHAP value) it removes all interactions with the other features, which prevents the possible leakage of patient information into the hospital attended value.

### 2.6 How much of the between-hospital thrombolysis use can be explained by the differences in the hospital identity and processes, and the differences in the patient populations?

The 10 features in the model can be classified into two subsets: 1) ‘patient descriptive features’ (features that describe the patients characteristics), and 2) ‘hospital descriptive features’ (features that describe the hospital’s identity or process). To analyse the influence that each subset of features has on the thromoblysis rate, we calculated the ‘subset SHAP value’ for each feature, which only includes the components of its SHAP value that contain effects from the features in the same subset. This is expressed as the sum of the main effect and the interaction effects with the other features in the same subset. For each subset of features (hospital, or patient) we fitted a multiple regression to predict the hospitals observed thrombolysis rate from the mean subset SHAP value of each feature, for patients attending each hospital (using values from the *all data* model). We also fitted a multiple regression using the subset SHAP values for all 10 features (both hospital and patient descriptive features). For this analysis, we only included the single one-hot encoded feature for the attended hospital.

### 2.7 Variation in hospital thrombolysis use for patient subgroups

Informed by the SHAP values, we analysed the observed and predicted use of thrombolysis in eleven subgroups of patients: one subgroup for ‘ideally’ thrombolysable patients, nine ‘sub-optimal’ thrombolysable patient subgroups (one subgroup per feature), and one subgroup with two sub-optimal features. We based the ideally thrombolysable definition on observing the relationships between feature values and thrombolysis use. The ‘sub-optimal’ patient subgroups were selected from all patients, apart from using a cut-off of one feature for each group. The eleven patient subgroups are defined as:

1. An *‘ideally’* thrombolysable patient:

- Mid-level stroke severity (NIHSS in range 10-25)
- Short arrival-to-scan time (less than 30 minutes)
- Stroke caused by infarction
- Precise stroke onset time known
- No pre-stroke disability (mRS 0)
- Not taking atrial fibrillation anticoagulants
- Short onset-to-arrival time (less than 90 minutes)
- Younger than 80 years old
- Onset not during sleep
2. Patients with a mild stroke severity (NIHSS less than 5)
3. Patients where stroke onset time known imprecisely
4. Patients with pre-stroke disability (mRS greater than 2)
5. Patients with a haemorrhagic stroke
6. Patients with 60-90 minutes arrival-to-scan time
7. Patients with use of AF anticoagulants
8. Patients with 150-180 minutes onset-to-arrival time
9. Patients with onset during sleep
10. Patients aged over 80 years old
11. Patients with a mild stroke severity (NIHSS less than 5) and with estimated stroke onset time

The observed thrombolysis use at each hospital was taken from the SSNAP dataset, with data limited to the patients that matched the patient characteristics and attending each hospital (and so the patient population was different for each hospital).

In order to reveal the variation in thrombolysis that was due to hospital decision-making we predicted thrombolysis use for the same patient subgroups at each hospital by using the *10k holdout* model.

## 3 Results

### 3.1 Variation in observed hospital thrombolysis use

Thrombolysis use in the original data varied between hospitals from 1.5% to 24.3% of all patients, and 7.3% to 49.7% of patients arriving within 4 hours of known (precise or estimated) stroke onset.

### 3.2 Feature selection

The best model with 1, 2, 5, 10, 25 & all 60 original features had ROC AUCs of 0.715, 0.792, 0.891, 0.919, 0.923 & 0.922. We selected 10 features for all subsequent work, which were:

- *Arrival-to-scan time*: Time from arrival at hospital to scan (minutes)
- *Infarction*: Stroke type (1 = infarction, 0 = haemorrhage)
- *Stroke severity*: National Institutes of Health Stroke Scale (NIHSS) score on arrival
- *Precise onset time*: Onset time (1 = precise, 0 = best estimate)
- *Prior disability level*: Disability level (modified Rankin Scale; mRS) before stroke
- *Stroke team*: Stroke team attended (hospital identifier)
- *Use of anticoagulants*: Use of prior anticoagulant (1 = Yes, 0 = No)
- *Onset-to-arrival time*: Time from onset of stroke to arrival at hospital (mins)
- *Onset during sleep*: Did stroke occur in sleep? (1 = Yes, 0 = No)
- *Age*: Age (as midpoint of 5 year age bands)

Correlations between the 10 features were measured using coefficients of determination (r-squared). All r-squared were less than 0.05 except a) age and prior disability level (r-squared 0.146), and b) onset during sleep and precise onset time (r-squared 0.078).

#### 3.2.1 Model accuracy

Model accuracy was measured using stratified 5-fold cross validation. Overall accuracy was 85.0% (83.9% sensitivity and specificity could be achieved simultaneously). The ROC AUC was 0.918. The model predicted hospital thrombolysis use at each hospital with very good accuracy (r-squared = 0.977, figure 1). This maintains the high accuracy of our previously published model with overall accuracy of 84.3% (83.2% sensitivity and specificity could be achieved simultaneously), and mean ROC AUC of 0.906^14^.

**Figure 1:**
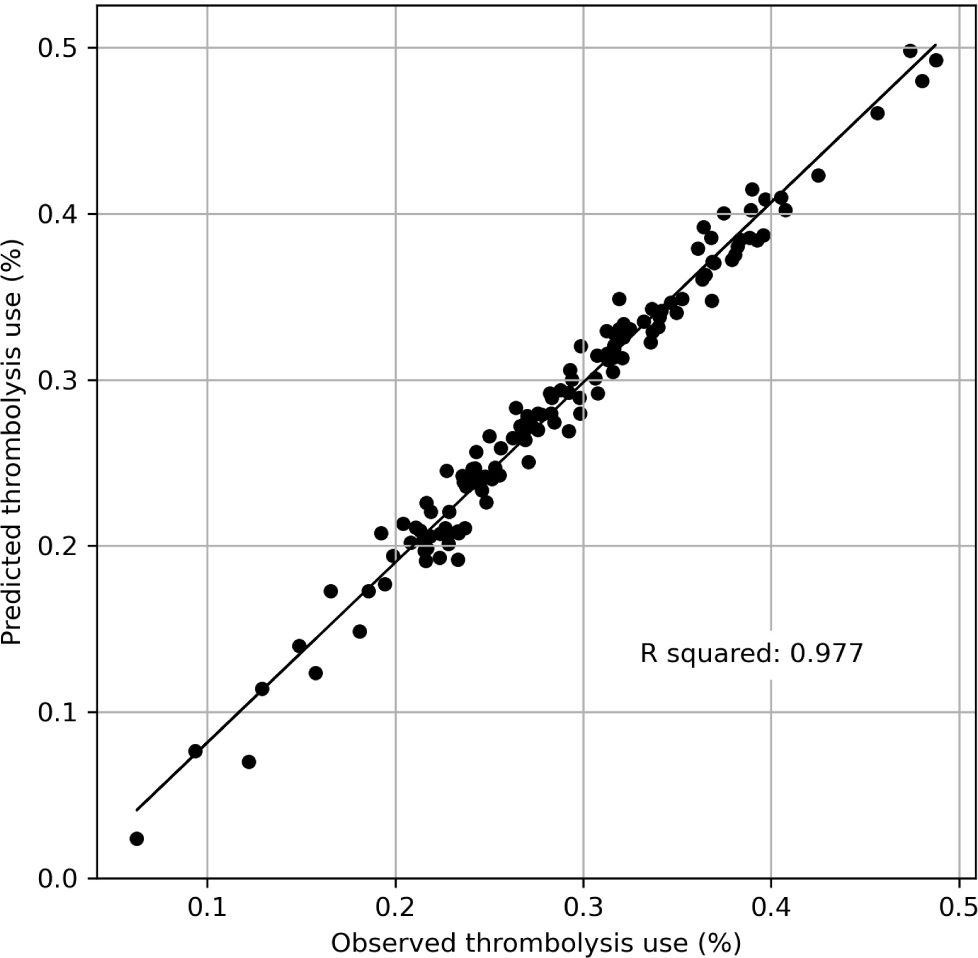
Comparison of predicted and observed thrombolysis use for 132 hospitals.

The appendix contains further model accuracy analysis.

### 3.3 Individual patient SHAP values

SHAP values are presented as how they affect log odds of receiving thrombolysis, but for individual predictions, probability values are more intuitive. Figure 2 shows waterfall plots for an example of a patient with low (top) and high (bottom) probability of receiving thrombolysis. Waterfall plots show the influence of features for an individual prediction (in our case, patient). The SHAP model starts with a base prediction of a 24% probability of receiving thrombolysis, before feature values are taken into account. For the patient with a low probability of receiving thrombolysis, the two most influential features reducing the probability of receiving thrombolysis were a long arrival-to-scan time (138 minutes) and a low stroke severity (NIHSS=2). For the patient with a high probability of receiving thrombolysis, the two most influential features increasing the probability of receiving thrombolysis were a short arrival-to-scan time (17 minutes) and a moderate stroke severity (NIHSS=14).

**Figure 2:**
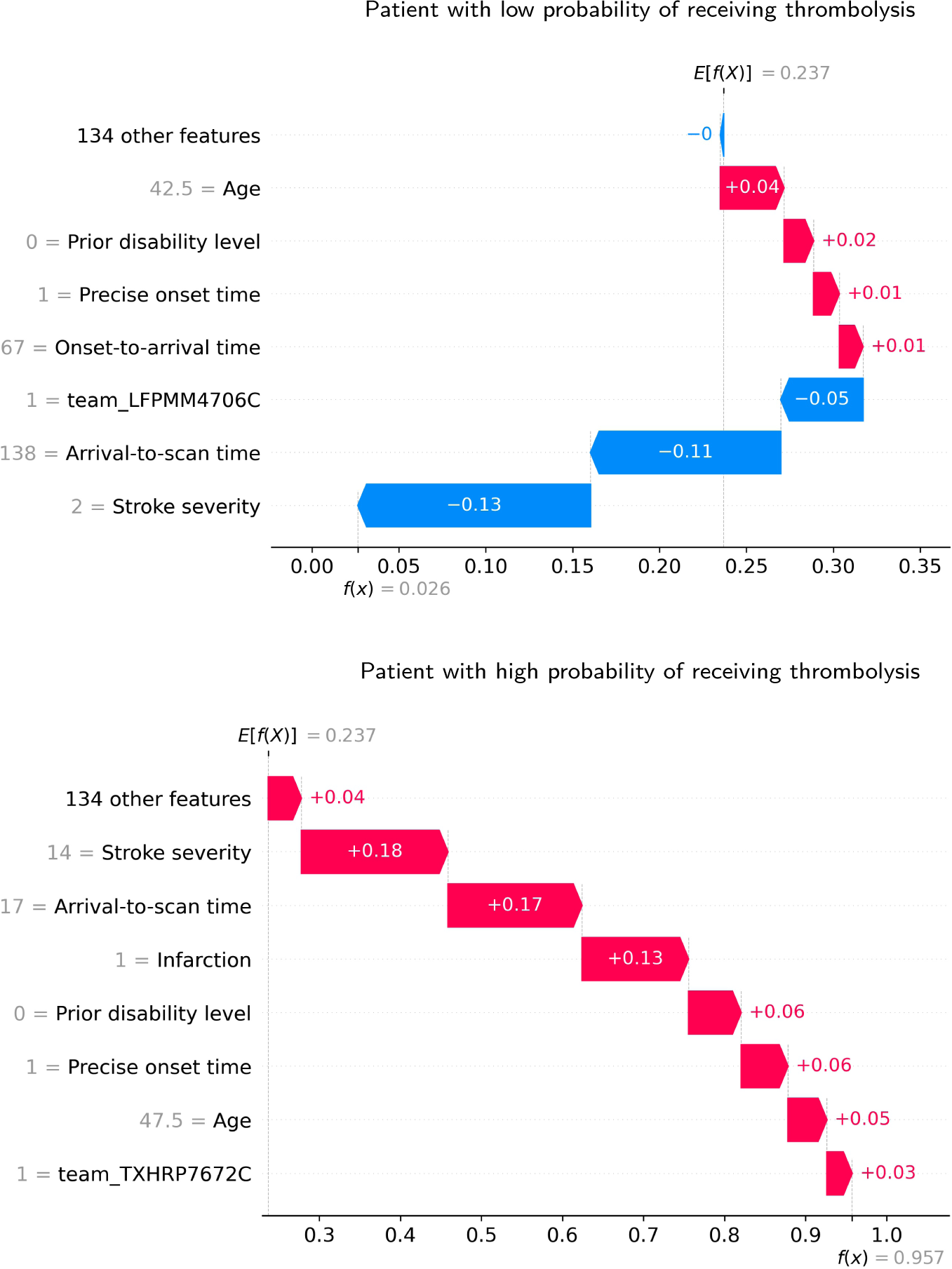
Waterfall plots showing the influence of each feature on the predicted probability of a single patient receiving thrombolysis. Top: An example of a patient with a low probability (2.6%) of receiving thrombolysis. Bottom: An example of a patient with a high probability (95.7%) of receiving thrombolysis.

### 3.4 The relationship between feature values and the odds of receiving thrombolysis

Figure 3 shows the relationship between patient level feature values and their SHAP values. Key observations are (with SHAP influence converted from log-odds to odds):

- *Stroke type*: The SHAP values for stroke type show that the model effectively eliminated any probability of receiving thrombolysis for non-ischaemic (haemorrhagic) stroke, with the odds of receiving thrombolysis falling by over 6,000 fold.
- *Arrival-to-scan time*: The odds of receiving thrombolysis reduced by about 9 fold over the first 120 minutes of arrival-to-scan time.
- *Stroke severity (NIHSS)*: The odds of receiving thrombolysis were lowest at NIHSS 0, increased and peaked at NIHSS 15-25, and then fell again with higher stroke severity (NIHSS above 25). The difference between minimum odds (at NIHSS 0) and maximum odds (at 15-25) of receiving thrombolysis was 25-30 fold.
- *Stroke onset time type (precise vs. estimated)*: The odds of receiving thrombolysis were about 3 fold greater for precise onset time than estimated onset time.
- *Disability level (mRS) before stroke*: The odds of receiving thrombolysis fell about 6 fold between mRS 0 and 5.
- *Use of AF anticoagulants*: The odds of receiving thrombolysis were about 5 fold greater for no use.
- *Onset-to-arrival time*: The odds of receiving thrombolysis were similar below 120 minutes, then fell about 3 fold between 120 and 240 minutes.
- *Age*: The odds of receiving thrombolysis were similar below 80 years old, then fell about 2 fold between 80 and 110 years old.
- *Onset during sleep*: The odds of receiving thrombolysis were about 4 fold lower for onset during sleep.
- *Hospital attended*: There was a 13 fold difference in odds of receiving thrombolysis between hospitals.

**Figure 3:**
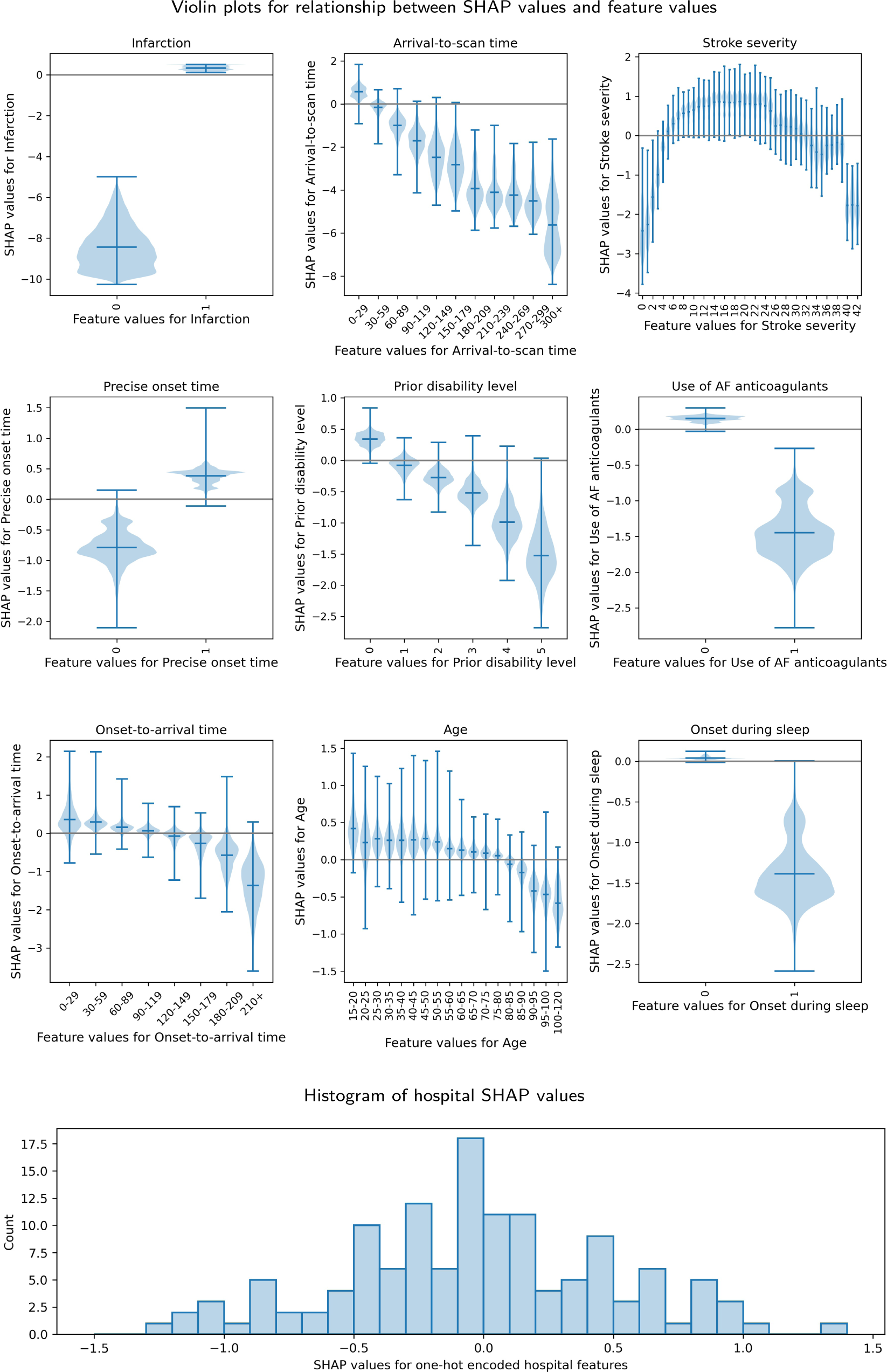
Plots showing the relationship between SHAP values and feature values. Top: Violin plots showing the relationship between SHAP values and feature values. The horizontal line shows the median SHAP value. The plots are ordered in ranked feature importance (using the mean absolute SHAP value across all instances). Bottom: Histogram showing the frequency of SHAP values for the one-hot encoded hospital feature.

In summary the most thrombolysable patients have: an infarction stroke type; shorter arrival-to-scan times; moderate to severe stroke severity with NIHSS 10-25; a precise onset time; lower pre-stroke disability; were not taking anticoagulant medication; shorter onset-to-arrival times; were younger; did not have onset during sleep; attend a hospital with a high predisposition to use thrombolysis.

### 3.5 How much of the between-hospital thrombolysis use can be explained by the identity of the hospital attended?

The mean hospital attended SHAP main effect value correlated with the observed hospital thrombolysis rate with an r-squared of 0.558 (figure 4, left), suggesting that 56% (P<0.0001) of the between-hospital variance in thrombolysis use may be explained by the attended hospitals’ SHAP main effect values, i.e. the hospitals’ predisposition and/or preparedness to use thrombolysis.

**Figure 4:**
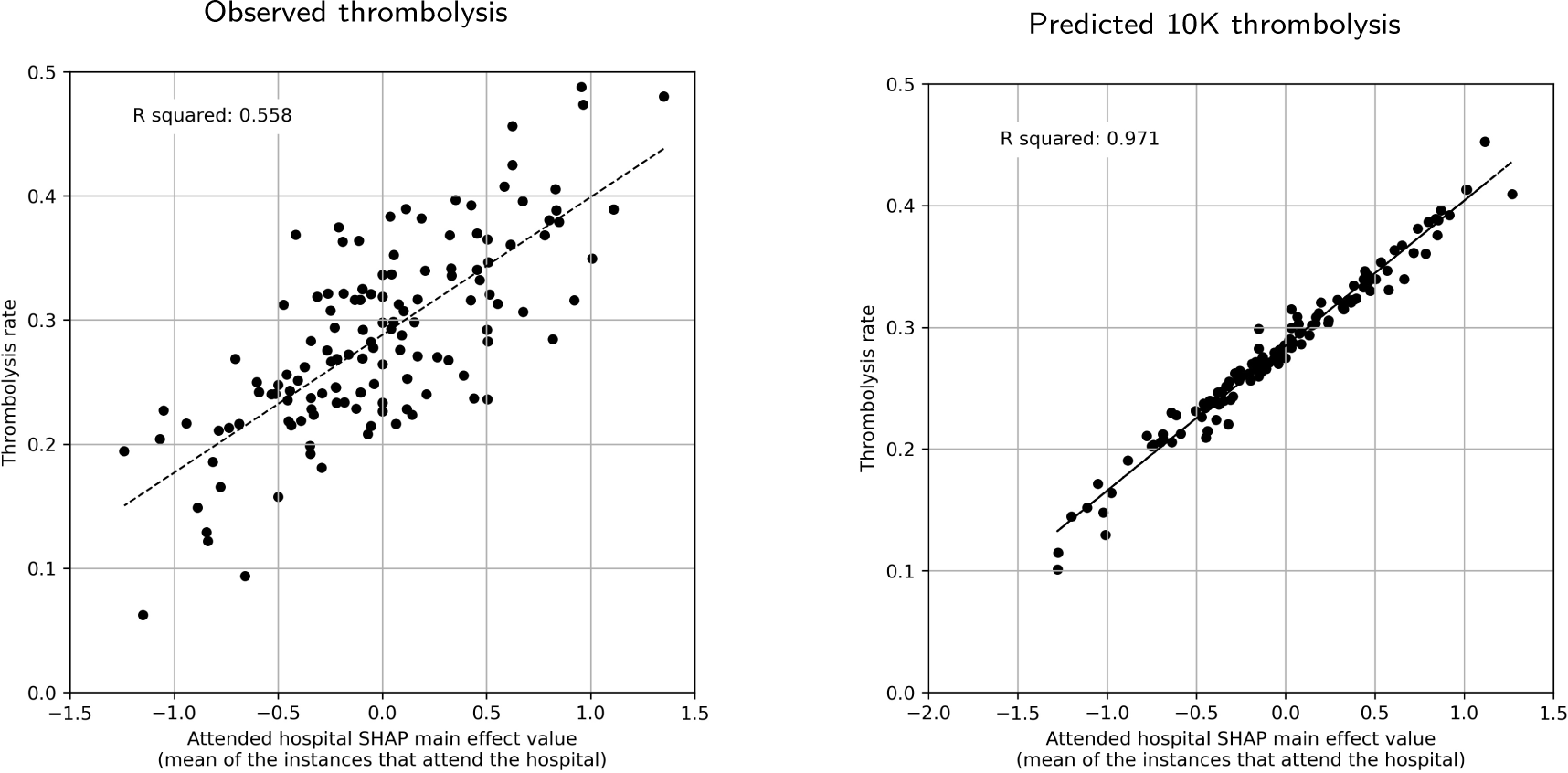
Correlations between hospital attended SHAP main effect value and the observed thrombolysis use at each hospital. Left: Observed thrombolysis (using the all data model). Right: Predicted 10k cohort thrombolysis rate (using the 10k holdout model).

Using the *10k holdout model*, the predicted use of thrombolysis across the 132 hospitals for the identical 10k cohort of patients (not used in training the model) ranged from 10% to 45%. The mean hospital attended SHAP main effect value for the 10k cohort of patients correlated very closely with the predicted thrombolysis use in the 10k cohort of patients at each hospital, with an r-squared of 0.971 (figure 4, right), confirming that the hospital attended SHAP main effect value is providing a direct insight into a hospitals’ propensity to use thrombolysis.

### 3.6 How much of the between-hospital thrombolysis use can be explained by the differences in the hospital identity and processes, and the differences in the patient populations?

We predicted thrombolysis use using mean subset SHAP values for patients attending each hospital. Figure 5 shows that 36% (P<0.0001) of the variance in observed between-hospital thrombolysis use can be explained by the patient population, 74% (P<0.0001) can be explained by hospital identity and processes, and that 95% (P<0.0001) can be explained by the combined information from both the patient population and hospital identity and processes.

**Figure 5:**
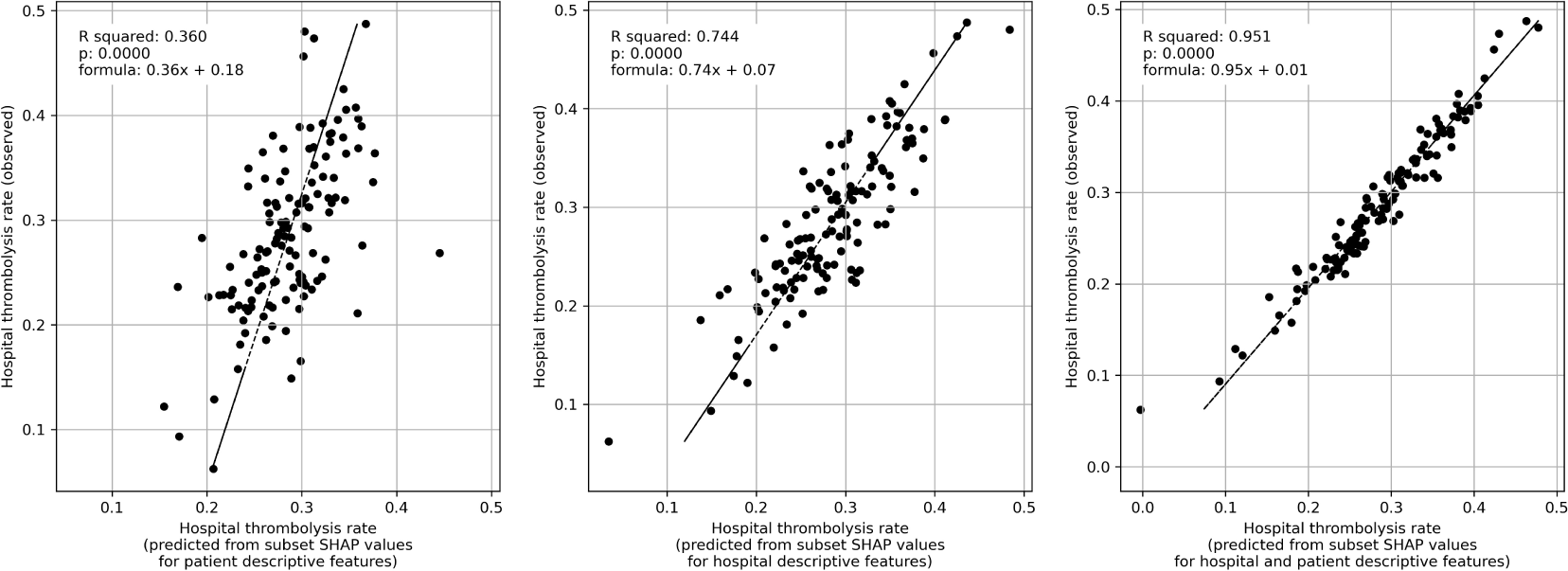
Multiple regression of subset SHAP values (mean of patients attending hospital) with hospital observed thrombolysis rate. Left: Subset SHAP values for the eight patient descriptive features (age, stroke severity, prior disability, onset-to-arrival time, stroke type, type of onset time, anticoagulants, and onset during sleep). Middle: Subset SHAP values for the two hospital descriptive features (arrival-to-scan time, and hospital attended). Right: Subset SHAP values for all 10 features (for both hospital and patient descriptive features).

### 3.7 Variation in hospital thrombolysis use for patient subgroups

Figure 6 shows boxplots of observed and predicted use of thrombolysis, broken down by patient subgroup. The nine subgroups of patients with one defined non-ideal feature (haemorrhagic stroke, arrival-to-scan time 60-90 mins, mild stroke severity, estimated stroke onset time, some pre-stroke disability, use of anticoagulants, onset-to-arrival time 150-180 mins, over 80 years old, and onset during sleep) all had reduced thrombolysis use than the complete patient population, and combining these non-ideal features reduced thrombolysis use further (illustrated by the patient subgroup with a mild stroke severity and an estimated stroke onset time). There was, however, significant variation between hospitals in use of thrombolysis in each of these subgroups. The observed and predicted thrombolysis use show the same general results, but some small differences existed: 1) The use of thrombolysis in *ideal* patients is a little lower in the observed vs predicted results (mean hospital thrombolysis use = 89% vs 99%), 2) The predicted results show a stronger effect of combining non-ideal features, 3) The observed thrombolysis rate shows higher between-hospital variation than the predicted thrombolysis rate. This may be partly explained by the observed thrombolysis rate being based on different patients at each hospital, but may also be partly explained by actual use of thrombolysis being slightly more variable than predicted thrombolysis use (which will follow general hospital patterns, and will not include, for example, between-clinician variation at each hospital).

**Figure 6:**
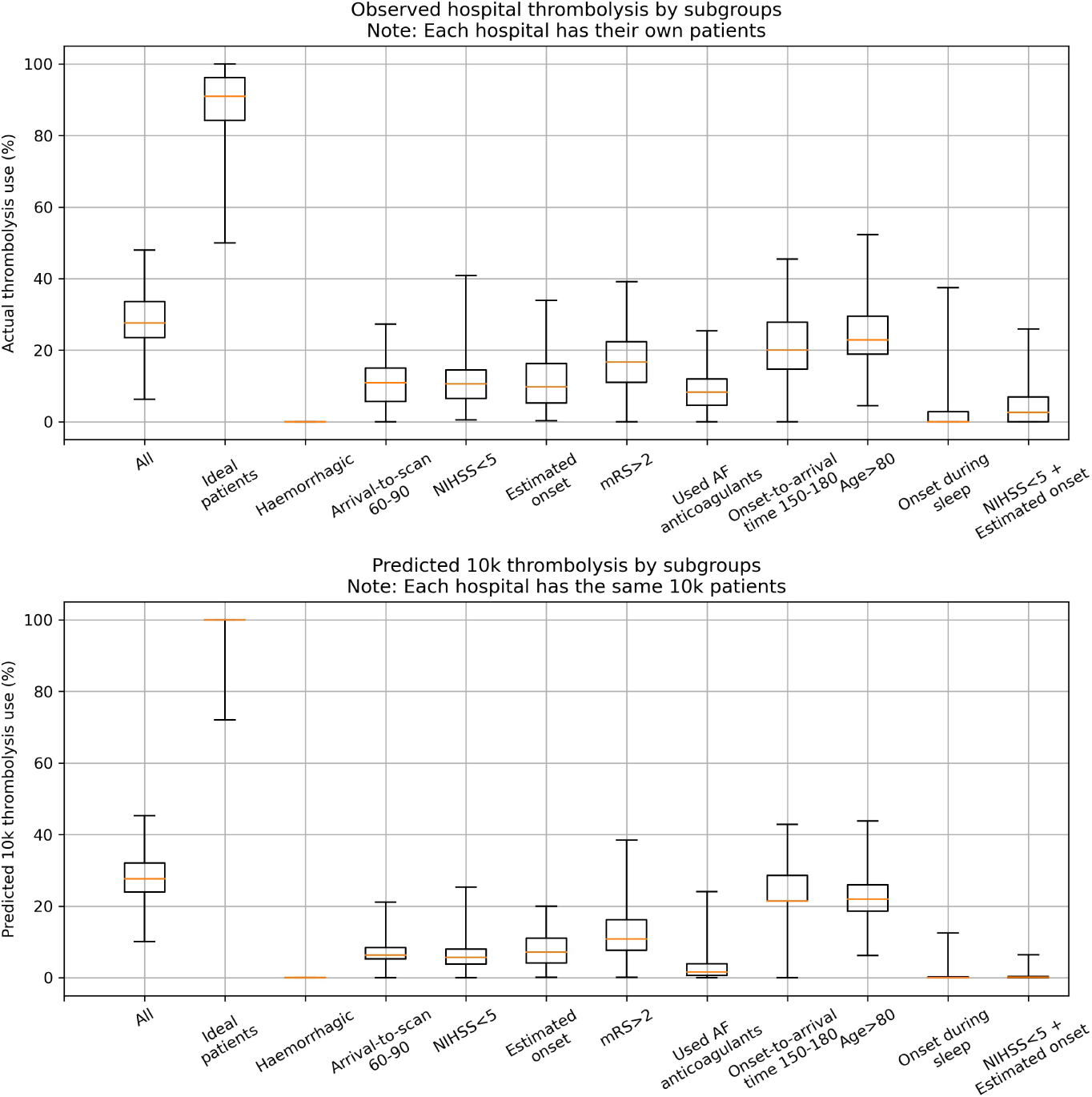
Boxplot for either observed (top) or predicted (bottom) use of thrombolysis for subgroups of patients. The ‘ideal patients’ subgroup has a mid-level stroke severity (NIHSS in range 10-25), short arrival-to-scan time (less than 30 minutes), stroke caused by infarction, precise stroke onset time known, no pre-stroke disability (mRS 0), not taking any atrial fibrillation anticoagulants, short onset-to-arrival time (less than 90 minutes), younger than 80 years old, and onset not during sleep. The single features are ordered in ranked feature importance (using the mean absolute SHAP value across all instances).

## 4 Discussion

We have built on our previous work to predict thrombolysis use from patient level data, by creating an *explainable machine learning model* which maintains the high accuracy that we previously achieved (85%) ^14^. The model was well-calibrated, with the average predicted probability of thrombolysis for any group of patients matching the proportion of those patients who did receive thrombolysis. The SSNAP registry data used therefore appears to contain most of the information used to make thrombolysis decisions in clinical practice. To further support this, when we identified a group of patients that the model predicts would receive thrombolysis all the time in the large majority of hospitals, the observed thrombolysis rate of that group was 90% in half the hospitals.

We have not designed our model to indicate whether an individual patient *should* receive thrombolysis. Despite our high accuracy, there will be relevant patient details not known to us that could preclude use of thrombolysis. Rather we have used machine learning to determine the usual patterns of use of thrombolysis that are common across hospitals, and vary between hospitals.

In general, using SHAP values to uncover the relationship between patient characteristics and the probability of receiving thrombolysis, we found that the probability of receiving thrombolysis fell with increasing arrival-to-scan times, was dependent on stroke severity with the probability of receiving thrombolysis being highest between NIHSS 10 and 25, was lower when onset time was estimated rather than known precisely, and fell sharply with increasing disability prior to stroke. These patterns are similar to the observations of a discrete choice experiment with hypothetical patients ^12^, but in our study we confirm these patterns in actual use of thrombolysis, can be quantitative about the effect, and we add the importance of time-to-scan and whether an onset time is known precisely. Using SHAP we can identify the general effect of each patient characteristic, such as stroke severity, and then also whether individual hospitals are influenced to a greater or lesser extent by that characteristic.

Hospital attended SHAP main effect values correlated very closely with the predicted use of thrombolysis in a 10k cohort of patients, confirming that the hospital attended SHAP main effect value provides a measure of the predisposition of a hospital to use thrombolysis. We found that hospital processes explained 74% of the variance in observed thrombolysis, suggesting that clinical decision-making around thrombolysis accounts for the majority of variance in use of thrombolysis between hospitals for those patients who arrive at hospital within 4 hours of known (precise or estimated) stroke onset time.

After observing the general patterns that exist in the use of thrombolysis, we created a subgroup of patients reflecting what appeared to be an *ideal* candidate for thrombolysis, and also a subgroup per feature where we expected to see lower use of thrombolysis (haemorrhage, longer arrival to scan, low stroke severity, estimated onset time, pre-stroke disability, using anticoagulants, longer onset to arrival, older age, and onset during sleep). Observed thrombolysis in these groups reflected the patterns identified by the SHAP analysis. For the *ideal* candidates of thrombolysis, half of stroke units would give thrombolysis to at least 90% of these patients, but some units gave it to significantly fewer patients. Use of thrombolysis in the other subgroups of patients was, as expected, lower, but use also varied significantly between hospitals, with use ranging between 0% and 50% for these subgroups of less ideal patients. These patterns, of lower but varying use, were repeated with expected use of thrombolysis in the same 10k patient cohort of patients.

A patient with acute ischaemic stroke presenting with a non-ideal feature will cause their likelihood to receive thrombolysis to be reduced. Hospitals, however, vary in how they respond to this information. The predisposition of a hospital to give thrombolysis, revealed in the SHAP value for each hospital, determines the level of acceptance they have to non-ideal features for their decision making to still be in favour of thrombolysis: a high predisposition gives them more tolerance to absorb a larger number, or higher magnitude, of non-ideal features.

This novel analysis examines and aids understanding of between-hospital variation in clinical decision making in the acute stroke setting. The application of this method is not restricted to stroke, or to England and Wales. It can be applied to any clinical decision where there is routinely collected data that quantifies the patients pathway up to the point of the decision, and the features recorded represent a sufficient quantity of the information that clinicians use to make their decision.

The use of large datasets such as SSNAP to understand sources of variation in clinical practice between large number of acute stroke centres across the UK presents a unique opportunity to understand the specific influences behind the significant residual between-hospital variation in thrombolysis use. In particular, it allows national quality improvement projects such as SSNAP to counter one of the most common objections raised to comparative audit: that the patients presenting to any one particular site are in some way unique, thereby accounting for most of the variation in clinical quality between that site and all the others – what, informly, has been termed the ‘special hospital fallacy’. Although the patient population does vary between hospitals, and will contribute to the thrombolysis use achievable by an individual hospital (being able to explain 34% of the variance in between-hospital thrombolysis use), the majority (74%) of between-hospital variation can be explained by hospital-level rather than patient-level factors. Another strength of the dataset is its richness. When the observed thrombolysis rate at each hospital is compared to the predicted thrombolysis rate, less than 3% unexplained variation remains – suggesting that no more than 3% of variation in thrombolysis use can be accounted for by factors that are not already measured in the national registry. SSNAP appears to contain the data that is needed for understanding variation in thrombolysis decisions.

It is disappointing that even though this disability-saving treatment was first licensed for approval for use over 20 years ago, it is still subject to such large 7-fold variation in clinical judgement or opinion regarding the selection of patients most appropriate for use. In our previous work ^13^, we have shown that increasing the uptake of thrombolysis through the administration of treatment to more patients and sooner after stroke, offers the prospect of more than doubling the proportion of patients after stroke who are left with little or no disability (mRS 0 or 1). At a time when there is an appropriate focus of effort on expanding the use of endovascular therapy in acute ischaemic stroke, it is sobering to consider how much population benefit there still remains to accrue from the fullest possible implementation of a cheaper technology that has been available for over 20 years. Far greater scrutiny of such residual variation in clinical practice is clearly warranted, given the extent to which it appears to be acting as a barrier to successful implementation. Recent studies have highlighted that clinicians can be reluctant to modify their behaviour in response to audit and feedback when it is not seen to be clinically meaningful, recent or reliable ^24^, so the full potential of audit and feedback is not realised ^25^ despite the evidence of a beneficial effect especially when baseline performance is low ^26^. The development of bespoke, individualised feedback (at least at hospital level) based on actual and recent activity may increase the impact of efforts at data-driven quality improvement targeted at increasing overall uptake of thrombolysis through reducing variation.

### 4.1 Limitations

This machine learning study is necessarily limited to data collected for the national stroke audit. Though we have high accuracy, and can identify clear patterns of use of thrombolysis, the data will not be sufficient to provide a decision-support tool. Nor is it a causal model. We may also be missing information that could otherwise have improved the accuracy still further. We also necessarily analyse thrombolysis at hospital level rather than at the level of the individual clinician. The model has high accuracy and can identify clear patterns, suggesting the capability to identify and characterise a centre’s culture in the use of thrombolysis, but we do not identify variation in thrombolysis between individual clinicians in the same hospital.

SHAP is a popular method for explaining the predictions of machine learning models, but it does have some limitations. SHAP values are an approximation of the Shapley values, from which they are based, in order to calculate them efficiently. Even so, SHAP can be computationally expensive and slow for large datasets and complex models. SHAP can only help to explain the fitted model, and so it can only be as good as the model (with caveats around training data containing bias, or incomplete information). Another limitation is the interpretation of SHAP values and its components (the main effect and interactions) can be challenging. To aid our dissemination of the findings from SHAP we have engaged with clinicians, patients and carers to learn how best to communicate this information. SHAP assumes feature independence - that is the assumption that the values of each feature in a dataset are independent of the values of other features in the same dataset. In other words, the value of one feature should not have a direct effect on the value of another feature. We used feature selection to ensure that very little covariance existed between the 10 features that were included in the models.

We acknowledge that not all countries have a national stroke audit dataset, however we hope that this paper helps to demonstrate what type of analysis can be done should resources be allocated to collect their national data.

## Data Availability

Data cannot be not available.

## Acknowledgements

This research was funded by the National Institute for Health Research (NIHR), and supported by the NIHR Applied Research Collaboration (ARC) South West Peninsula. The views expressed are those of the authors and not necessarily those of the NHS, the NIHR or the Department of Health and Social Care.

We would like to thank the SAMueL project team (Lauren Asare, Leon Farmer, Julia Frost, Iain Lang, Kristin Liabo, Peter McMeekin, Keira Pratt-Boyden, Cathy Pope, Ken Stein, Penny Thompson) for their input into this work.

We would also like to thank our extended Patient and Carer Involvement team (David Burgess, Simon Douglas, Ian Hancock, Nicola Hancock, John Williams), and our expert advisory group (Ajay Bhalla, Gary Ford, Anthony Rudd, and Martin Utley).

## Data access and ethics

The NHS Health Research Authority decision tool^c^ was used to confirm that ethical approval was not required to access the data. No identifiable patient or hospital information were provided in the data, and anonymised hospital names were provided. Governance of the data and access to SSNAP was authorised by the Healthcare Quality Improvement Partnership^d^ (HQIP, reference HQIP303).

## Author approval

All authors have seen and approved this manuscript.

## Links to source code, online Jupyter book, and LaTeX paper

1. GitHub repository: https://github.com/samuel-book/samuel_shap_paper_122
2. Jupyter book: https://samuel-book.github.io/samuel_shap_paper_1/
3. LATEXpaper: https://github.com/samuel-book/overleaf_shap_paper_1_short

### Appendix

This work is licensed under a Creative Commons 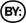 3.0 license.

#### A.1 Data

##### A.1.1 Data access

Data was obtained from the Sentinel Stroke National Audit (SSNAP^e^), managed through the Healthcare Quality Improvement Partnership (HQIP^f^). SSNAP has near-complete coverage of all acute stroke admissions in the UK (outside Scotland). All hospitals admitting acute stroke participate in the audit, and year-on-year comparison with Hospital Episode Statistics^g^ confirms estimated case ascertainment of 95% of coded cases of acute stroke.

The NHS Health Research Authority decision tool^h^ was used to confirm that ethical approval was not required to access the data. Data access was authorised by HQIP (reference HQIP303).

Data was retrieved for 246,676 emergency stroke admissions to acute stroke teams in England and Wales between 2016 and 2018 (three full years). 88,928 patients arrived within 4 hours of known stroke onset.

##### A.1.2 Data fields

###### Stroke Team

- *StrokeTeam*: Pseudonymised SSNAP ‘routinely admitting team’ unique identifier. For emergency care it is expected that each hospital has one stroke team (though post-72 hour care may be reported under a different team at that hospital).

###### Patient – general

- *Pathway*: Total number of team transfers, excluding community teams
- *S1AgeOnArrival*: Age on arrival aggregated to 5 year bands
- *MoreEqual80y*: Whether the patient is <= 80 years old at the moment of the stroke
- *S1Gender*: Gender
- *S1Ethnicity*: Patient Ethnicity. Aggregated to White, Black, Mixed, Asian and Other

###### Patient – pathway information

- *S1OnsetInHospital*: Whether the patient was already an inpatient at the time of stroke
- *S1OnsetToArrival min*: Time from symptom onset to arrival at hospital in minutes, where known and if out of hospital stroke
- *S1OnsetDateType*: Whether the date of onset given is precise, best estimate or if the stroke occurred while sleep
- *S1OnsetTimeType*: Whether the time of symptom onset given is precise, best estimate, not known
- *S1ArriveByAmbulance*: Whether the patient arrived by ambulance
- *S1AdmissionHour*: Hour of arrival, aggregates to 3 hour epochs
- *S1AdmissionDay*: Day of week at the moment of admission
- *S1AdmissionQuarter*: Year quarter (Q1: Jan-Mar; Q2:April-Jun; Q3: Jul-Sept; Q4: Oct-Dec)
- *S1AdmissionYear*: Year of admission
- *S2BrainImagingTime min*: Time from Clock Start to brain scan. In minutes. “Clock Start” is used throughout SSNAP reporting to refer to the date and time of arrival at first hospital for newly arrived patients, or to the date and time of symptom onset if patient already in hospital at the time of their stroke.
- *S2ThrombolysisTime min*: Time from Clock Start to thrombolysis. In minutes. “Clock Start” is used throughout SSNAP reporting to refer to the date and time of arrival at first hospital for newly arrived patients, or to the date and time of symptom onset if patient already in hospital at the time of their stroke.

###### Patient – comorbidities

- *CongestiveHeartFailure*: Pre-Stroke Congestive Heart Failure
- *Hypertension*: Pre-Stroke Systemic Hypertension
- *AtrialFibrillation*: Pre-Stroke Atrial Fibrillation (persistent, permanent, or paroxysmal)
- *Diabetes*: Comorbidities: Pre-Stroke Diabetes Mellitus
- *StrokeTIA*: Pre-Stroke history of stroke or Transient Ischaemic Attack (TIA)
- *AFAntiplatelet*: Only available if “Yes” to Atrial Fibrillation comorbidity. Whether the patient was on antiplatelet medication prior to admission
- *AFAnticoagulent*: Prior to 01-Dec-2017: Only available if “Yes” to Atrial Fibrillation comorbidity; From 01-Dec-2017: available even if patient is not in Atrial Fibrillation prior to admission. Whether the patient was on anticoagulant medication prior to admission
- *AFAnticoagulentVitK*: If the patient was receiving anticoagulant medication, was it vitamin K antagonists
- *AFAnticoagulentDOAC*: If the patient was receiving anticoagulant medication, was it direct oral anticoagulants (DOACs)
- *AFAnticoagulentHeparin*: If the patient was receiving anticoagulant medication, was it Heparin

###### Patient – NIH Stroke Scale

- *S2NihssArrival*: National Institutes of Health Stroke Scale score on arrival at hospital
- *BestGaze*: National Institutes of Health Stroke Scale Item 2 Best Gaze (higher values indicate more severe deficit)
- *BestLanguage*: National Institutes of Health Stroke Scale Item 9 Best Language (higher values indicate more severe deficit)
- *Dysarthria*: National Institutes of Health Stroke Scale Item 10 Dysarthria (higher values indicate more severe deficit)
- *ExtinctionInattention*: National Institutes of Health Stroke Scale Item 11 Extinction and Inattention (higher values indicate more severe deficit)
- *FacialPalsy*: National Institutes of Health Stroke Scale Item 4 Facial Paresis (higher values indicate more severe deficit)
- *LimbAtaxia*: National Institutes of Health Stroke Scale Item 7 Limb Ataxia (higher values indicate more severe deficit)
- *Loc*: National Institutes of Health Stroke Scale Item 1a Level of Consciousness (higher values indicate more severe deficit)
- *LocCommands*: National Institutes of Health Stroke Scale Item 1c Level of Consciousness Commands (higher values indicate more severe deficit)
- *LocQuestions*: National Institutes of Health Stroke Scale Item 1b Level of Consciousness Questions (higher values indicate more severe deficit)
- *MotorArmLeft*: National Institutes of Health Stroke Scale Item 5a Motor Arm - Left (higher values indicate more severe deficit)
- *MotorArmRight*: National Institutes of Health Stroke Scale Item 5b Motor Arm - Right (higher values indicate more severe deficit)
- *MotorLegLeft*: National Institutes of Health Stroke Scale Item 6a Motor Leg - Left (higher values indicate more severe deficit)
- *MotorLegRight*: National Institutes of Health Stroke Scale Item 6b Motor Leg - Right (higher values indicate more severe deficit)
- *Sensory*: National Institutes of Health Stroke Scale Item 8 Sensory (higher values indicate more severe deficit)
- *Visual*: National Institutes of Health Stroke Scale Item 3 Visual Fields (higher values indicate more severe deficit)

###### Patient – other clinical features

- *S2INR*: Patient’s International Normalised ratio (INR) on arrival at hospital (available since 01-Dec-2017)
- *S2INRHigh*: INR was greater than 10 on arrival at hospital (available since 01-Dec-2017)
- *S2INRNK*: INR not checked (available since 01-Dec-2017)
- *S2NewAFDiagnosis*: Whether a new diagnosis of Atrial Fibrillation was made on admission
- *S2RankinBeforeStroke*: Patient’s modified Rankin Scale score before this stroke (Higher values indicate more disability)
- *S2StrokeType*: Whether the stroke type was infarction or primary intracerebral haemorrhage
- *S2TIAInLastMonth*: Whether the patient had a Transient Ischaemic Attack during the last month. Item from the SSNAP comprehensive dataset questions (not mandatory)

###### Patient – thrombolysis given

- *S2Thrombolysis*: Whether the patient was given thrombolysis (clot busting medication)

###### Patient – reason stated for not giving thrombolysis

- *Age*: If the answer to thrombolysis given was “no but”, the reason was Age
- *Comorbidity*: If the answer to thrombolysis given was “no but”, the reason was Co-morbidity
- *Haemorrhagic*: If the answer to thrombolysis given was “no but”, the reason was Haemorrhagic stroke
- *Improving*: If the answer to thrombolysis given was “no but”, the reason was Symptoms Improving
- *Medication*: If the answer to thrombolysis given was “no but”, the reason was Medication
- *OtherMedical*: If the answer to thrombolysis given was “no but”, the reason was Other medical reason
- *Refusal*: If the answer to thrombolysis given was “no but”, the reason was Refusal
- *TimeUnknownWakeUp*: If the answer to thrombolysis given was “no but”, the reason was Symptom onset time unknown/wake-up stroke
- *TimeWindow*: If the answer to thrombolysis given was “no but”, the reason was Age
- *TooMildSevere*: If the answer to thrombolysis given was “no but”, the reason was Stroke too mild or too severe

#### A.2 Probability, odds, and Shap values (log odds shifts): A brief explanation

Many of us find it easiest to think of the chance of something occurring as a probability. For example, there might be a probability of 10% that it will rain today. That is the same as saying there will be one rainy day out of ten days for days with this given probability of rain.

In our stroke thrombolysis model, Shap values tell us how knowing something particular about a patient (such as the patient *feature*, ‘Is their stroke caused by a clot or a bleed?’) adjusts our prediction of whether they will receive thrombolysis or not.

This is made a little more complicated for us because Shap is usually reported as a *log odds shift*. It is useful for us to see how those relate to probabilities, and get a sense of how significant Shap values in the range of 0.5 to 5 (or −0.5 to −5) are, as that is a common range of Shap values that we will see in our models.

##### A.2.1 Probability

We will take the example that Shap reports that a model’s base probability prediction, before consideration of features is 0.25, or a 25% probability of receiving thrombolysis; that is 1 in 4 patients with this prediction would be expected to receive thrombolysis.

##### A.2.2 Odds

*Probability* expresses the chance of something happening as the number of positive occurrences as a fraction of all occurrences (i.e. the number of patients receiving thrombolysis as a fraction of the total number of patients).

*Odds* express the chance of something happening as the ratio of the number of positive occurrences (i.e. receiving thrombolysis) to the number of negative occurrences (i.e. *not* receiving thrombolysis).

If we have probability prediction of 0.25 would receive thrombolysis, that would mean 1 in 4 of those patients receive thrombolysis. Expressed as odds, for every one patient that receives thrombolysis, three will not. The odds are expressed as 1:3 or 1/3. This may also be calculated as a decimal (1 divided by 3), 0.333.

Odds (O) and probability (P) may be converted with the following equations:

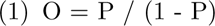

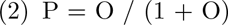

##### A.2.3 Shap values: Log odds shifts

Here we will calculate the effect of Shap values, and try and build some intuition on the size of effect Shap values of 0.5 to 5 give (we will look at positive and negative Shap values).

Shap usually outputs the effect of a particular feature in how much it shifts the odds. For reasons we will not go into here, that shift (which is the ‘Shap value’) is usually given in ‘log odds’ (the logarithm of the odds value). For the mathematically inclined, we use the natural log (*ln*).

Let’s look at some Shap values (log odds) and see how much they change the odds of receiving thrombolysis.

First we’ll look at the shift in odds the Shap values give. This is calculated as *shift = exp(Shap)* (table A.1).

**Table A.1:** The relationship between *odds* and *log odds*.

| SHAP (log odds) | Shift in odds (multiply original odds) |
| --- | --- |
| 0.5 | 1.65 |
| 1 | 2.72 |
| 2 | 7.39 |
| 3 | 20.1 |
| 4 | 54.6 |
| 5 | 148 |

###### Positive Shap values: a worked example

Now let us work through an example of starting with a known baseline *probability* (before we consider what we know about a particular patient feature), converting that to *odds*, applying a Shap *log odds shift* for that particular feature, and converting back to *probability* after we have applied the influence of that feature.

The the effects of those shifts on our baseline probability of 0.25 are shown in table A.2.

**Table A.2:** The effect of SHAP values between 0.5 and 5 on a base probability of 0.25

| Starting P | Starting O | SHAP | Shift (multiply O) | Shifted O | Shifted P (%) |
| --- | --- | --- | --- | --- | --- |
| 0.25 (25%) | 0.333 | 0.5 | 1.65 | 0.550 | 0.3547 (35.5%) |
| 0.25 (25%) | 0.333 | 1 | 2.72 | 0.907 | 0.4754 (47.5%) |
| 0.25 (25%) | 0.333 | 2 | 7.39 | 2.46 | 0.7112 (71.1%) |
| 0.25 (25%) | 0.333 | 3 | 20.1 | 6.70 | 0.8700 (87.0%) |
| 0.25 (25%) | 0.333 | 4 | 54.6 | 18.2 | 0.9479 (94.8%) |
| 0.25 (25%) | 0.333 | 5 | 148 | 49.5 | 0.9802 (98.0%) |

So, for example, a Shap value of 0.5 for one particular feature tells us that that particular feature in that patient shifts our expected probability of that patient receiving thrombolysis from 25% to 36%. A Shap value of 5 for the same feature would shift the probability of that patient receiving thrombolysis up to 98%.

###### Negative Shap values: a worked example

If we have a negative Shap value then odds are reduced (a Shap of −1 will lead to the odds being divided by 2.72, which is the same as multiplying by 1/2.72, which is 0.3679), as shown in table A.3.

**Table A.3:**
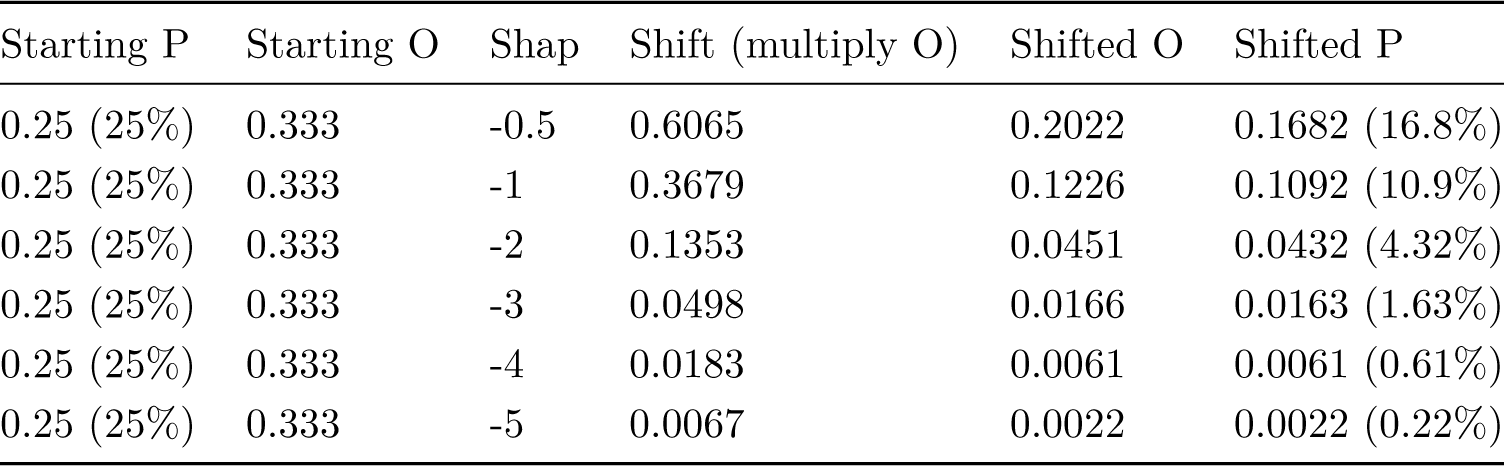
The effect of SHAP values between −0.5 and −5 on a base probability of 0.25

So, for example, a Shap value of −0.5 for one particular feature tells us that that particular feature in that patient shifts our expected probability of that patient receiving thrombolysis from 25% to 17%. A Shap value of 5 for the same feature would shift the probability of that patient receiving thrombolysis down to 2%.

##### A.2.4 Observations about Shap values

We begin to get some intuition on Shap values. A Shap value of 0.5 (or −0.5) leads to a small, but still noticeable, change in probability. Shap values of 5 or −5 have effectively pushed probabilities to one extreme or the other.

#### A.3 Variation in thrombolysis use

Thrombolysis use in the original data varied between hospitals (Figure A.7), from 1.5% to 24.3% of all patients, and 7.3% to 49.7% of patients arriving within 4 hours of known stroke onset.

**Figure A.7:**
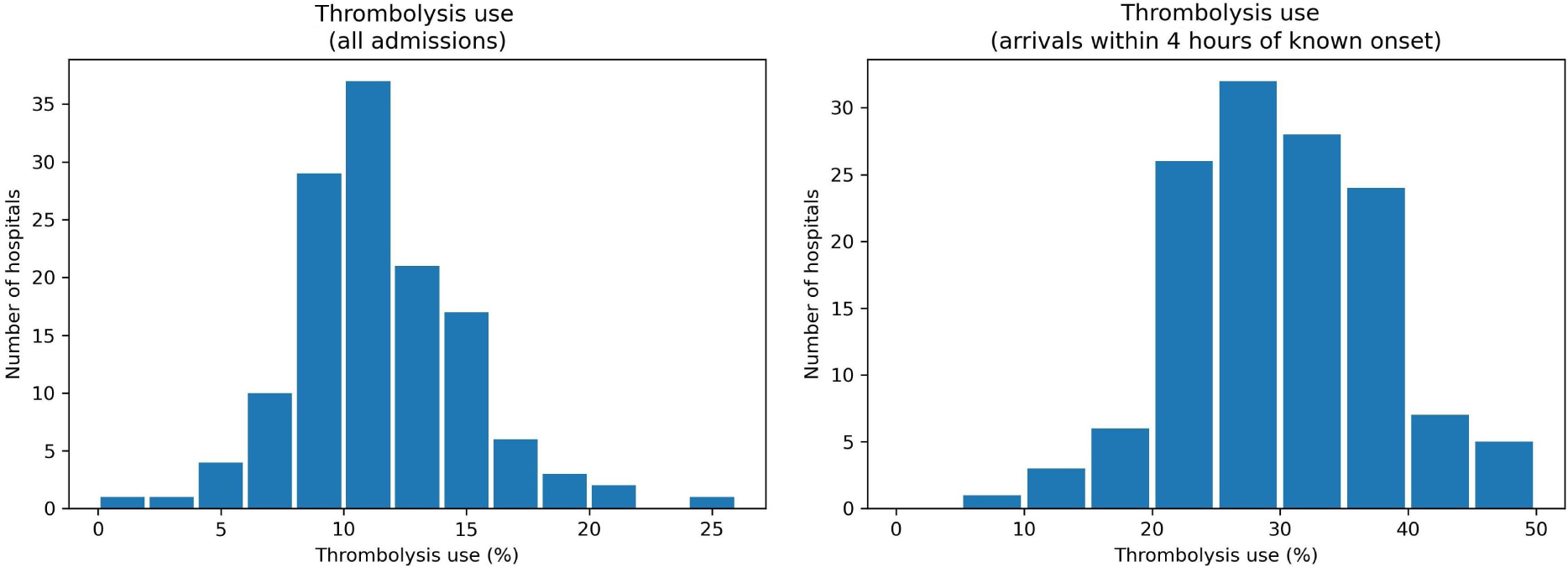
Histogram of observed thrombolysis use in 132 hospitals. Left: Thrombolysis shown as a percentage of all emergency stroke admissions. Right: Thrombolysis shown as a percentage of those patients who arrive at hospitals within 4 hours of known stroke onset.

#### A.4 Machine learning methods

All work was conducted in Python (v3.8). All code is available at:

1. GitHub repository: https://github.com/samuel-book/samuel_shap_paper_122
2. Jupyter book: https://samuel-book.github.io/samuel_shap_paper_1/

Our machine learning model used XGBoost (*eXtreme Gradient Boosting*, v1.5, https://pypi.org/ project/xgboost/). We used default settings apart from *learning rate* was set at 0.5 (see section A.11).

Machine learning models were explained using SHAP (*SHapley Additive exPlanations*, v0.41 https://pypi.org/project/shap/).

#### A.5 Feature selection

A simplified model was created by using *forward feature selection* where features were added in accordance to how much each one improved the Receiver Operating Characteristic (ROC) Area Under Curve (AUC). ROC AUC was measured using stratified k-fold validation (k=5). A model with all available 84 features had an ROC AUC of 0.922. A model with 10 features had an ROC AUC of 0.919.

The 10 features selected (Figure A.8) were:

- *Arrival-to-scan time*: Time from arrival at hospital to scan (mins)
- *Infarction*: Stroke type (1 = infarction, 0 = haemorrhage)
- *Stroke severity*: Stroke severity (NIHSS) on arrival
- *Precise onset time*: Onset time type (1 = precise, 0 = best estimate)
- *Prior disability level*: Disability level (modified Rankin Scale) before stroke
- *Stroke team*: Stroke team attended
- *Use of AF anticoagulants*: Use of atrial fibrillation anticoagulant (1 = Yes, 0 = No)
- *Onset-to-arrival time*: Time from onset of stroke to arrival at hospital (mins)
- *Onset during sleep*: Did stroke occur in sleep?
- *Age*: Age (as middle of 5 year age bands)

NOTE: All results from this point forward will use the 10 feature model

#### A.6 Correlations within the 10 selected features

Correlations between the 10 features were measured using coefficients of determination (r-squared). All r-squared were less than 0.15, and all r-squared were less than 0.05 except 1) age and prior disability level (r-squared 0.146), and 2) onset during sleep and precise onset time (r-squared 0.078). All correlations are shown in table A.4.

**Table A.4:** Correlations between the 10 features selected for the XGBoost machine learning model.

| Variable 1 | Variable 2 | r-squared |
| --- | --- | --- |
| Age | Prior disability level | 0.1462 |
| Onset during sleep | Precise onset time | 0.0784 |
| Stroke severity | Prior disability level | 0.0454 |
| Stroke severity | Infarction | 0.0386 |

| Variable 1 | Variable 2 | r-squared |
| --- | --- | --- |
| Precise onset time | Onset-to-arrival time | 0.0344 |
| Stroke severity | Age | 0.0268 |
| Age | Use of AF anticoagulants | 0.0207 |
| Stroke severity | Onset-to-arrival time | 0.0186 |
| Precise onset time | Prior disability level | 0.0131 |
| Age | Precise onset time | 0.0090 |
| Prior disability level | Use of AF anticoagulants | 0.0070 |
| Onset during sleep | Onset-to-arrival time | 0.0043 |
| Onset-to-arrival time | Age | 0.0038 |
| Use of AF anticoagulants | Infarction | 0.0033 |
| Prior disability level | Onset-to-arrival time | 0.0022 |
| Precise onset time | Arrival-to-scan time | 0.0021 |
| Use of AF anticoagulants | Stroke severity | 0.0019 |
| Arrival-to-scan time | Stroke severity | 0.0019 |
| Precise onset time | Use of AF anticoagulants | 0.0016 |
| Stroke severity | Onset during sleep | 0.0011 |
| Infarction | Onset-to-arrival time | 0.0007 |
| Infarction | Onset during sleep | 0.0007 |
| Infarction | Precise onset time | 0.0006 |
| Onset-to-arrival time | Arrival-to-scan time | 0.0004 |
| Arrival-to-scan time | Prior disability level | 0.0001 |
| Onset-to-arrival time | Use of AF anticoagulants | 0.0001 |
| Stroke severity | Precise onset time | 0.0000 |
| Arrival-to-scan time | Age | 0.0000 |
| Use of AF anticoagulants | Onset during sleep | 0.0000 |
| Prior disability level | Onset during sleep | 0.0000 |
| Infarction | Age | 0.0000 |
| Use of AF anticoagulants | Arrival-to-scan time | 0.0000 |
| Onset during sleep | Arrival-to-scan time | 0.0000 |
| Arrival-to-scan time | Infarction | 0.0000 |
| Age | Onset during sleep | 0.0000 |
| Prior disability level | Infarction | 0.0000 |

#### A.7 Model accuracy

Model accuracy was measured using stratified 5-fold cross validation. The key results are shown in table A.5.

**Figure A.8:**
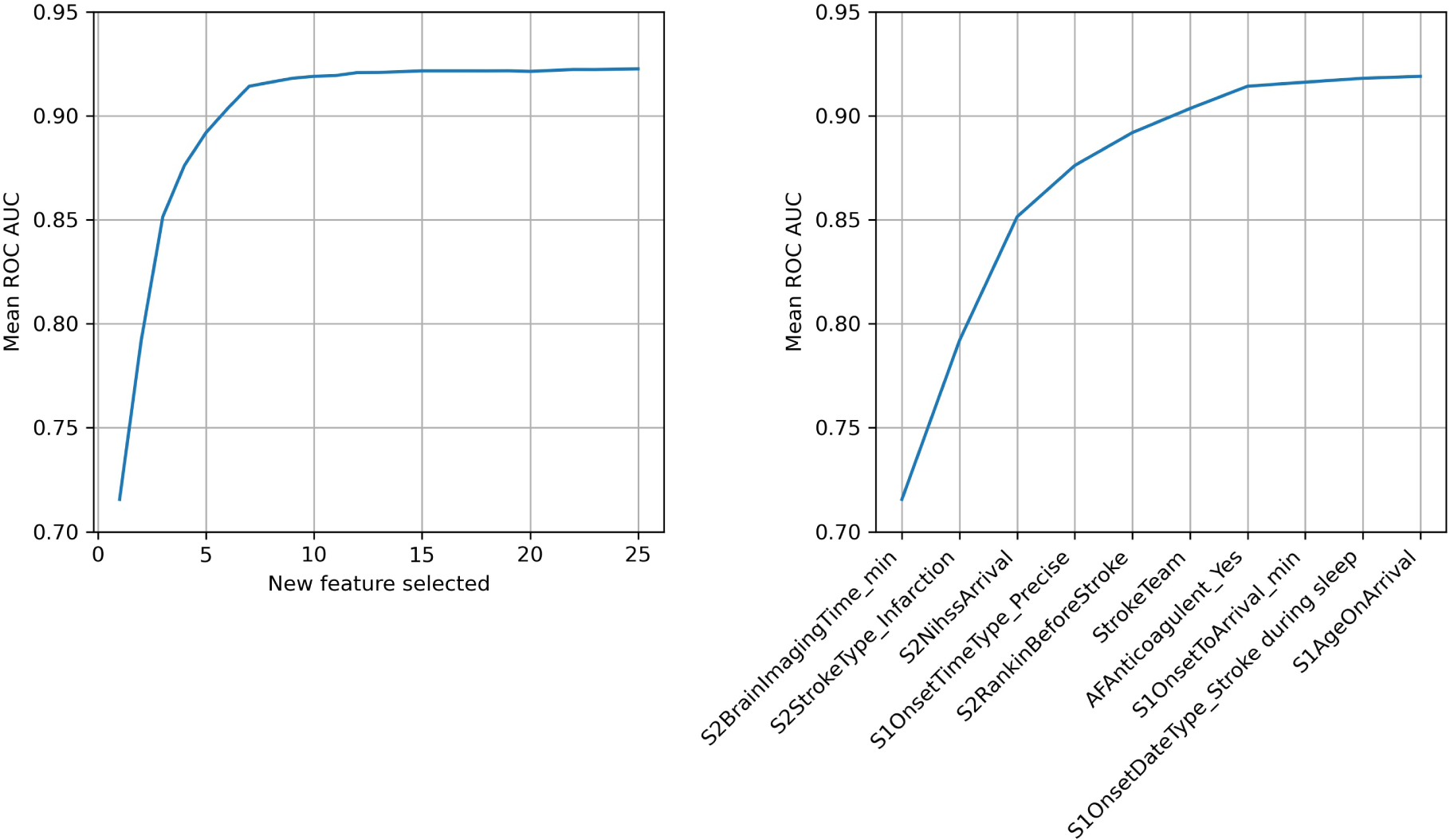
The effect of increasing the number of features on model accuracy measured by Receiver Operating Characteristic (ROC) Area Under Curve (AUC). Left: Improvement with ROC AUC with selection of up to 25 features. Right: Improvement with ROC AUC with selection of the best 10 features. ROC was measured with stratified 5-fold cross-validation. Results show the mean of the 5-fold replicates.

**Table A.5:** Accuracy of 10 feature XGBoost model in predicting thrombolysis use in patients arriving at hospital within 4 hours of known stroke onset.

| Accuracy measurement | mean | std |
| --- | --- | --- |
| Actual positive rate | 0.296 | 0.000 |
| Actual negative rate | 0.704 | 0.000 |
| Predicted positive rate | 0.294 | 0.002 |
| Predicted negative rate | 0.706 | 0.002 |
| Accuracy | 0.850 | 0.004 |
| Sensitivity (recall) | 0.743 | 0.004 |
| Specificity | 0.894 | 0.004 |
| Precision | 0.747 | 0.007 |
| ROC AUC | 0.918 | 0.003 |
| Balanced sensitivity/specificity | 0.839 | 0.003 |

We found an overall accuracy of 85.0%, with a balanced accuracy. The predicted thrombolysis rate of 29.4% was very close to the observed thrombolysis rate of 29.6%.

Figure A.9 shows the receiver operating characteristic curve, along with the trade-off between sensitivity and specificity.

**Figure A.9:**
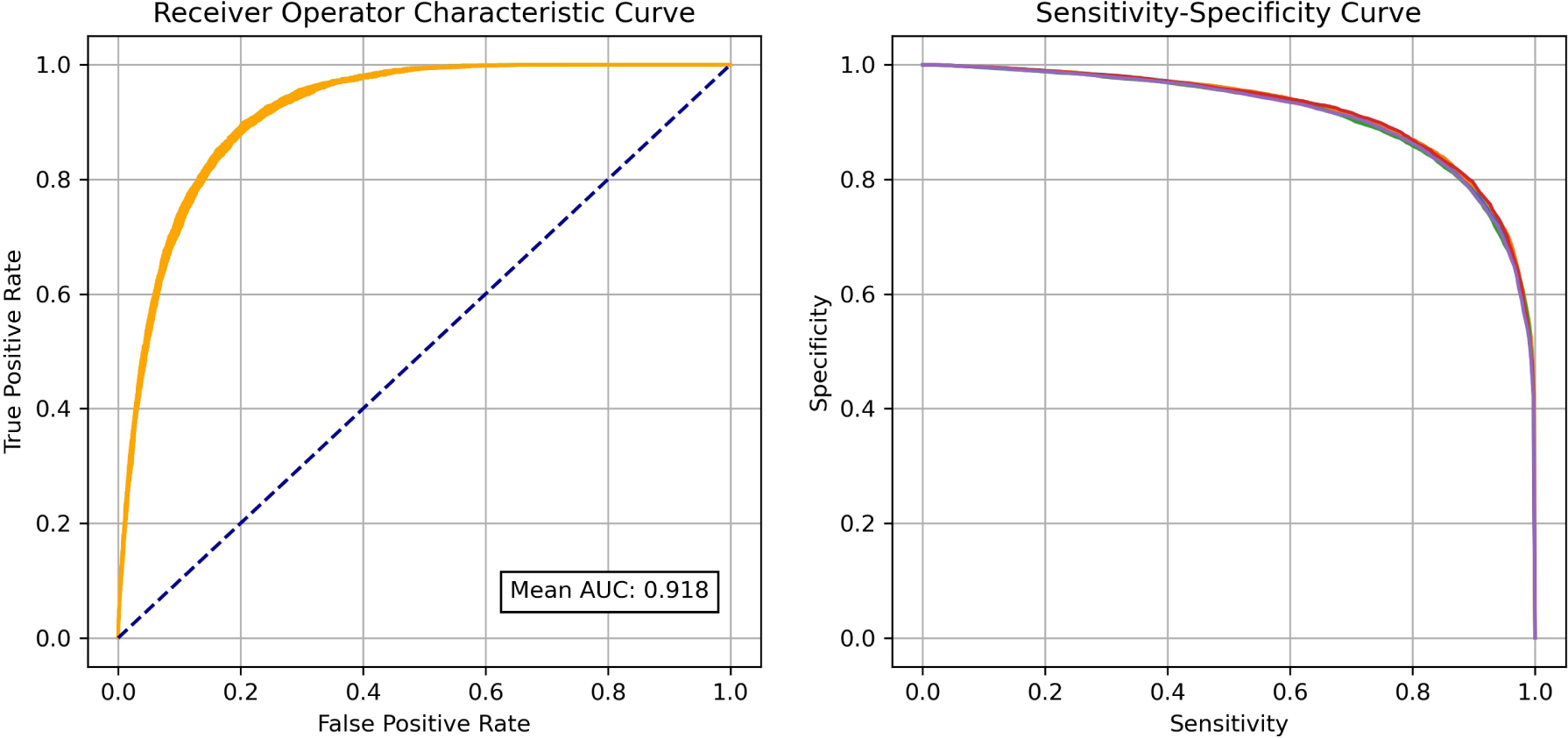
Model accuracy of a XGBoost model using 10 features. Left: Receiver Operating Characteristic (ROC) Area Under Curve (AUC). Right: The trade-off between Sensitivity and Specificity. Accuracy was measured with stratified 5-fold cross-validation, and both charts show all 5 k-fold replicates.

#### A.8 Model calibration

The model calibration was checked by binning predictions by probability, and comparing the mean predicted probability with the fraction that were actually positive (table A.6 and figure A.10). In a well-calibrated model, in each bin the average probability of receiving thrombolysis should be close to the proportion of patients who actually received thrombolysis. Results demonstrated that the model was naturally well-calibrated, and was not in need of any calibration correction. As expected, the fraction of predictions that were correct is related to the predicted probability of receiving thrombolysis (when predictions were close to 50% probability of receiving thrombolysis the model was correct about 50% of the time, whereas when the model had predictions of less than 10% or greater than 90% probability of receiving thrombolysis, the model was be correct about 90% of the time).

Nearly 50% of patients fell in the 0-10% probability of receiving thrombolysis - that is the model gave a confident prediction that the these patients would not receive thrombolysis, with the model being correct in these predictions 98% of the time.

**Figure A.10:**
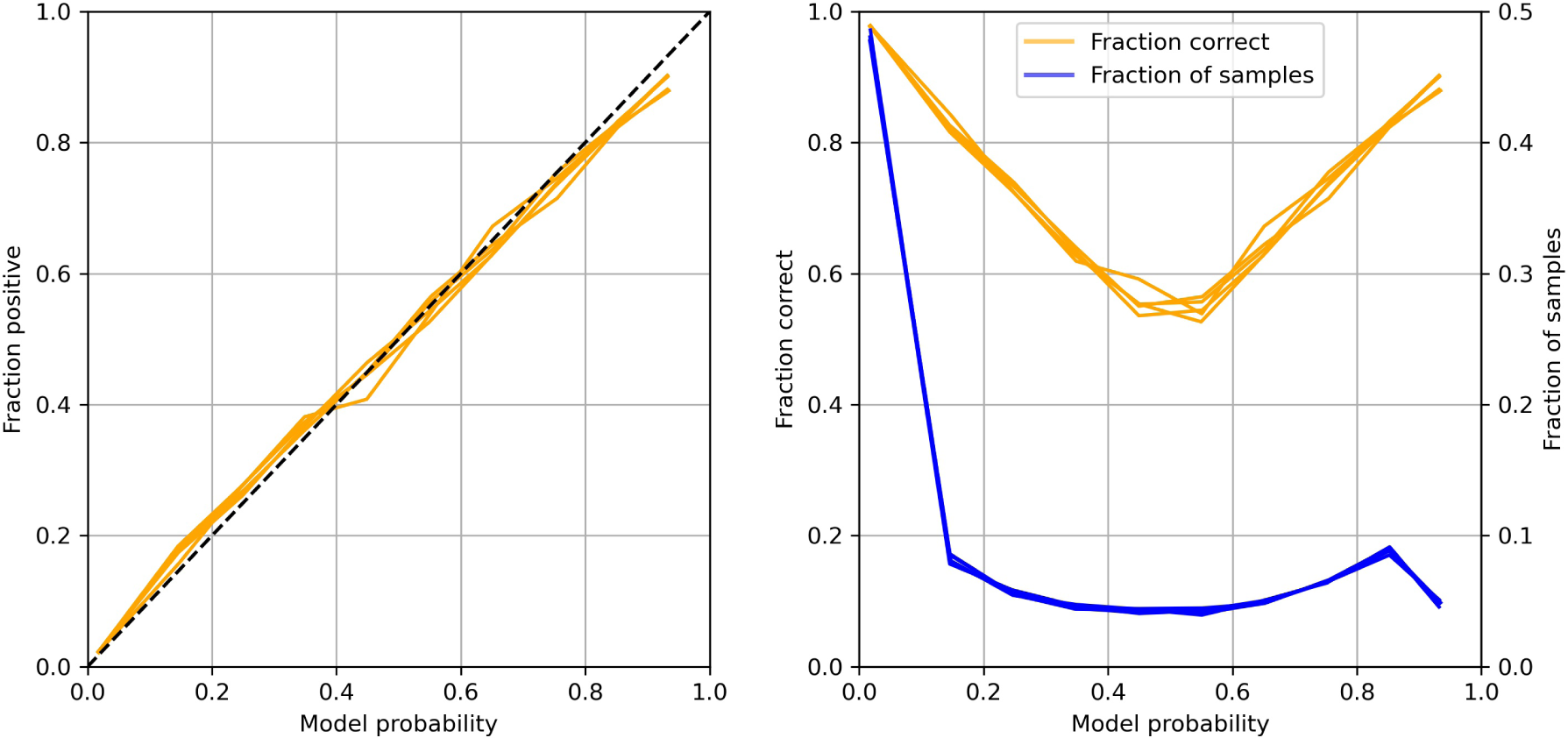
Calibration check of the model. Left: The proportion of patients receiving thrombolysis for binned probability of receiving thrombolysis. Right: The proportion of predictions in each bin (blue), and the proportion of predictions that are correct (orange). Plot show results for all 5 k-fold replicates.

**Table A.6:**
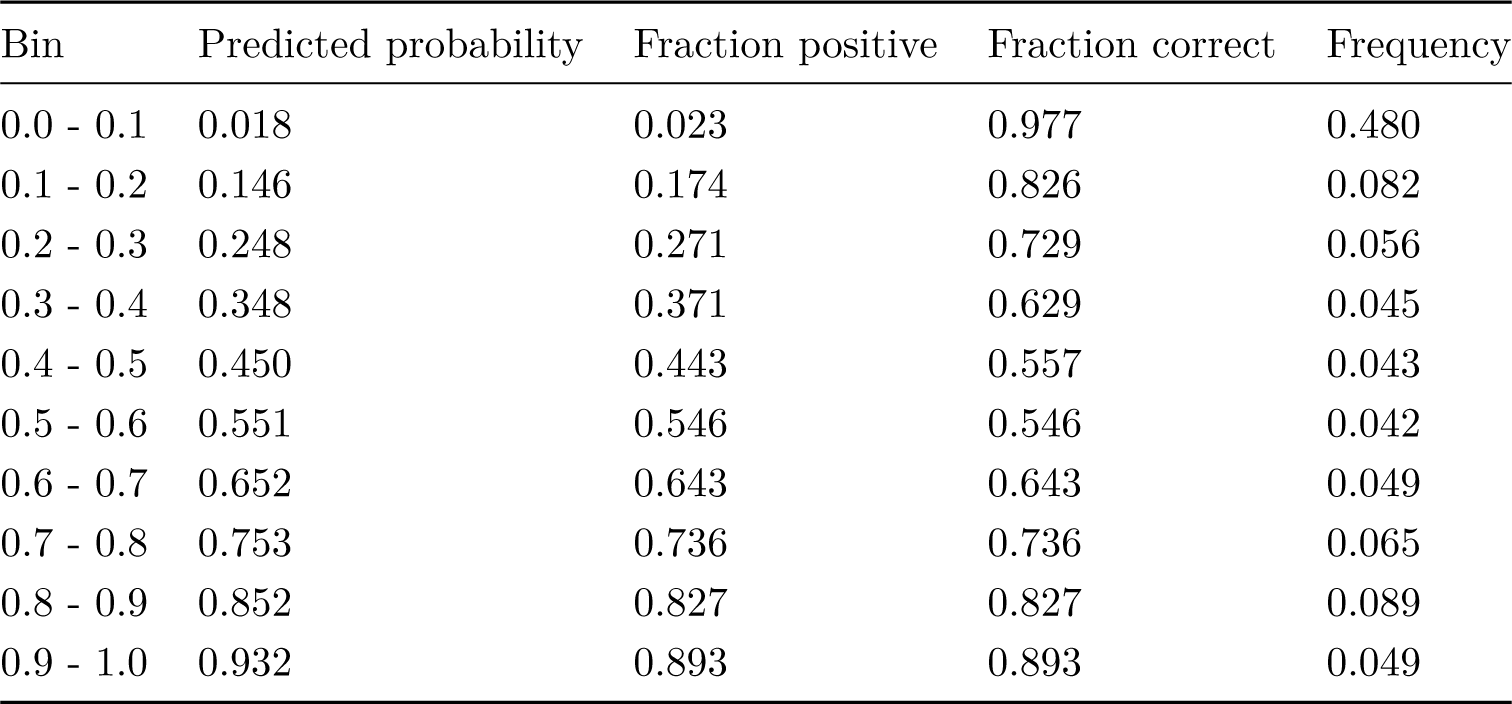
Model calibration based on binning by predicted probability of thrombolysis.

#### A.9 Evaluating variation in model predictions and predicted 10k cohort thrombolysis rate using bootstrap models

Data was split into a training set of 78,928 patients, and a test set of 10k patients. 30 models were trained, each with a different bootstrap sample of the training set and with a different model random seed. For each of the 10k test set, we evaluated the variation in the predicted probability of receiving thrombolysis (figure A.11). The mean of these standard deviations was 0.057, but the variation depended on the probability, with variation peaking at about 0.13 when the prediction probability of receiving thrombolysis was around 0.5.

Additionally, we used the models and test set to predict thrombolysis use at each of the 132 hospitals if the 10k cohort of patients had attended each of the hospitals (by changing the hospital one-hot encoding, figure A.12). We predicted the thrombolysis use at each hospital, and examined the variation between the 30 bootstrapped models. The mean of the standard deviation of bootstrap replicates was 1.7% (where hospital thrombolysis use rates were 10% to 45%).

**Figure A.11:**
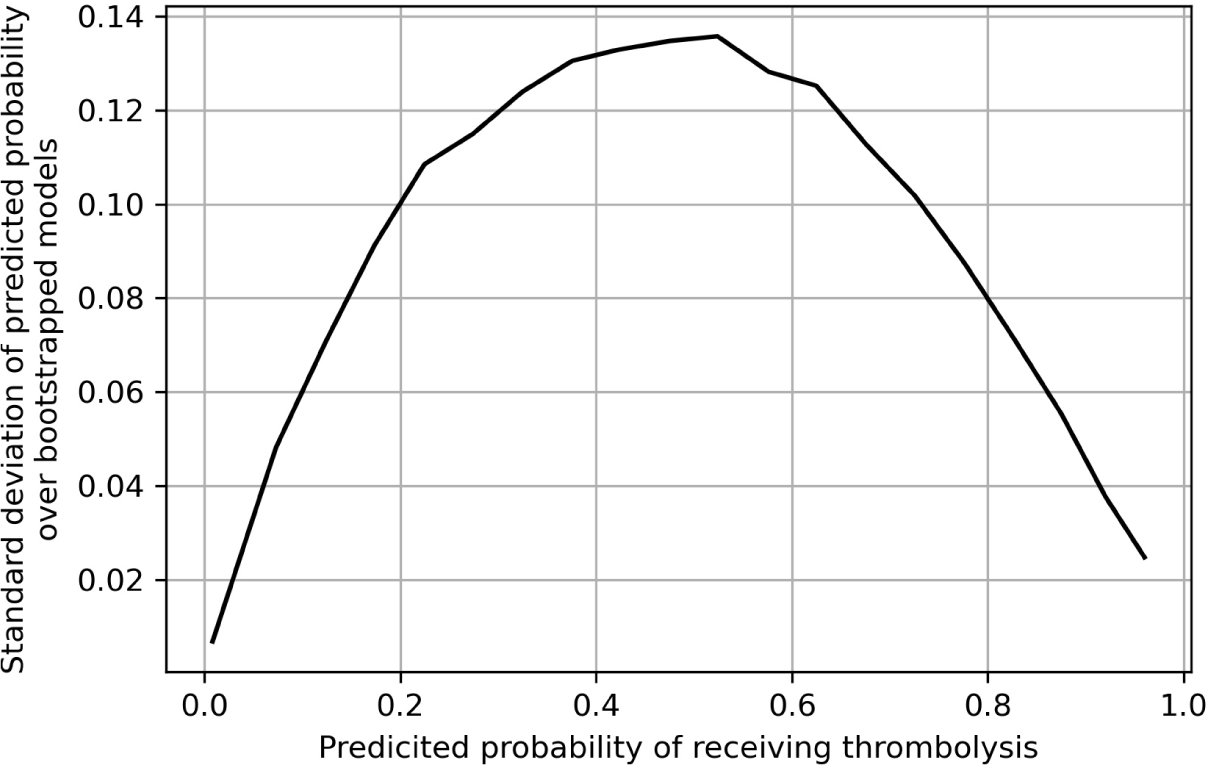
Standard deviation of predicted probability of receiving thrombolysis, from 30 bootstrapped models predicting the probability of receiving thrombolysis in 10k patients. Results are binned by predicted probability.

**Figure A.12:**
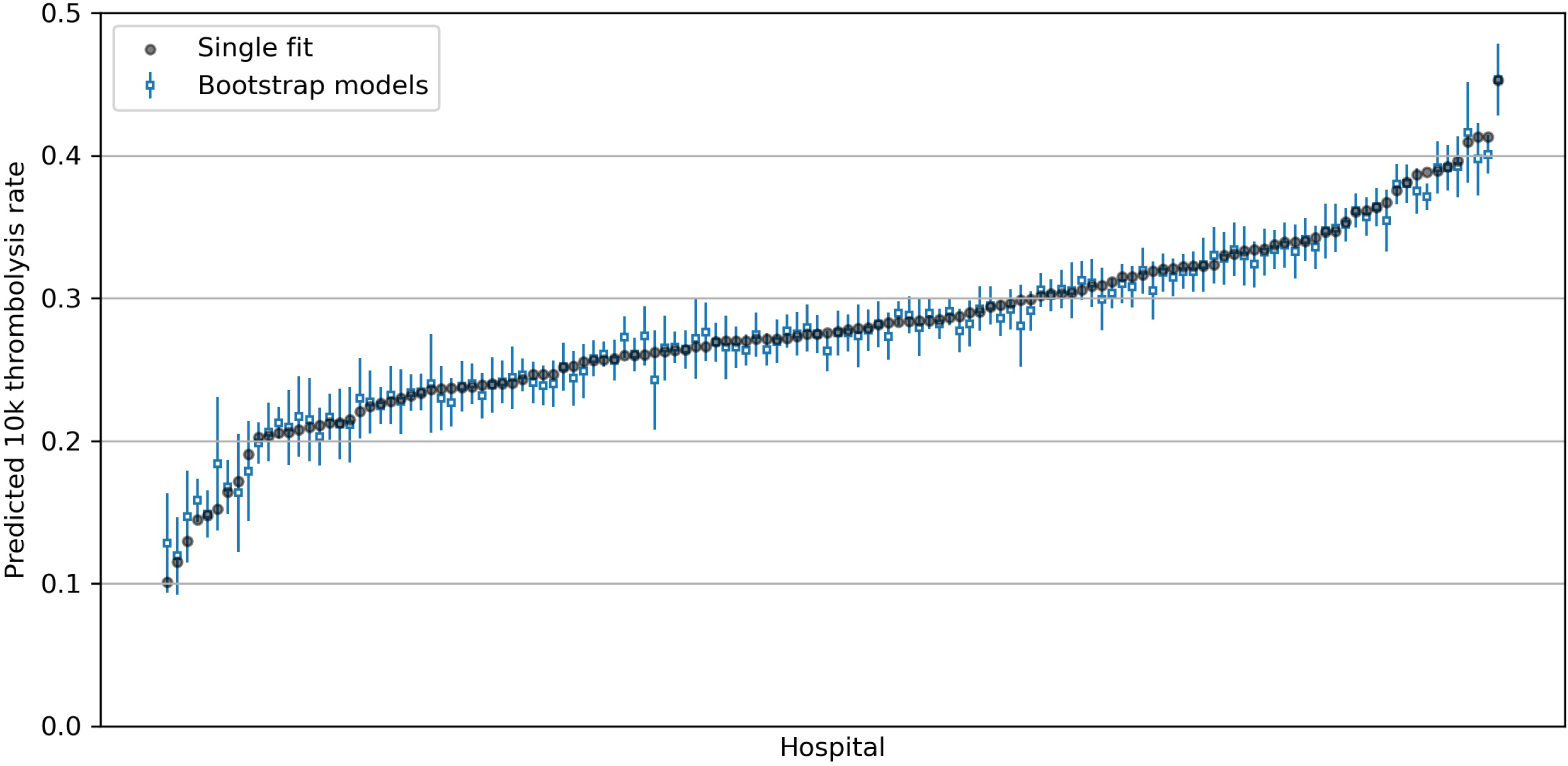
Mean and standard deviation of predicted thrombolysis use at 132 hospitals from 30 bootstrapped models. Results are for predicted thrombolysis use for the same 10k patient cohort for each hospital. In addition to the results for the bagging models, the predicted thrombolysis use for a single model with bootstrap sampling is shown. Results are ordered by thrombolysis use at each hospital predicted from the single non-bootstrap model.

Bagging experiments were repeated with *Baysian Bootstrapping* based on weighting training samples using a Dirichlet distribution. Very similar results were achieved, with a mean standard deviation of bootstrap replicate probability predictions of 0.054, and a mean standard deviation of bootstrap replicate 10k thrombolysis use in hospitals of 1.6%.

The evaluation of bootstrapped replicates gave us confidence that a single model fit would be sufficient.

#### A.10 Learning curves

Learning curves evaluate the relationship between training set size and model accuracy. Learning curves were performed using stratified 5-fold validation, and by random sampling (without replacement) of the training set (figure A.13. The maximum accuracy achieved was 85% using 70k training instances, 82.5% accuracy was achieved with 4k training instances. There was a shallow improvement between 4k and 70k training points.

**Figure A.13:**
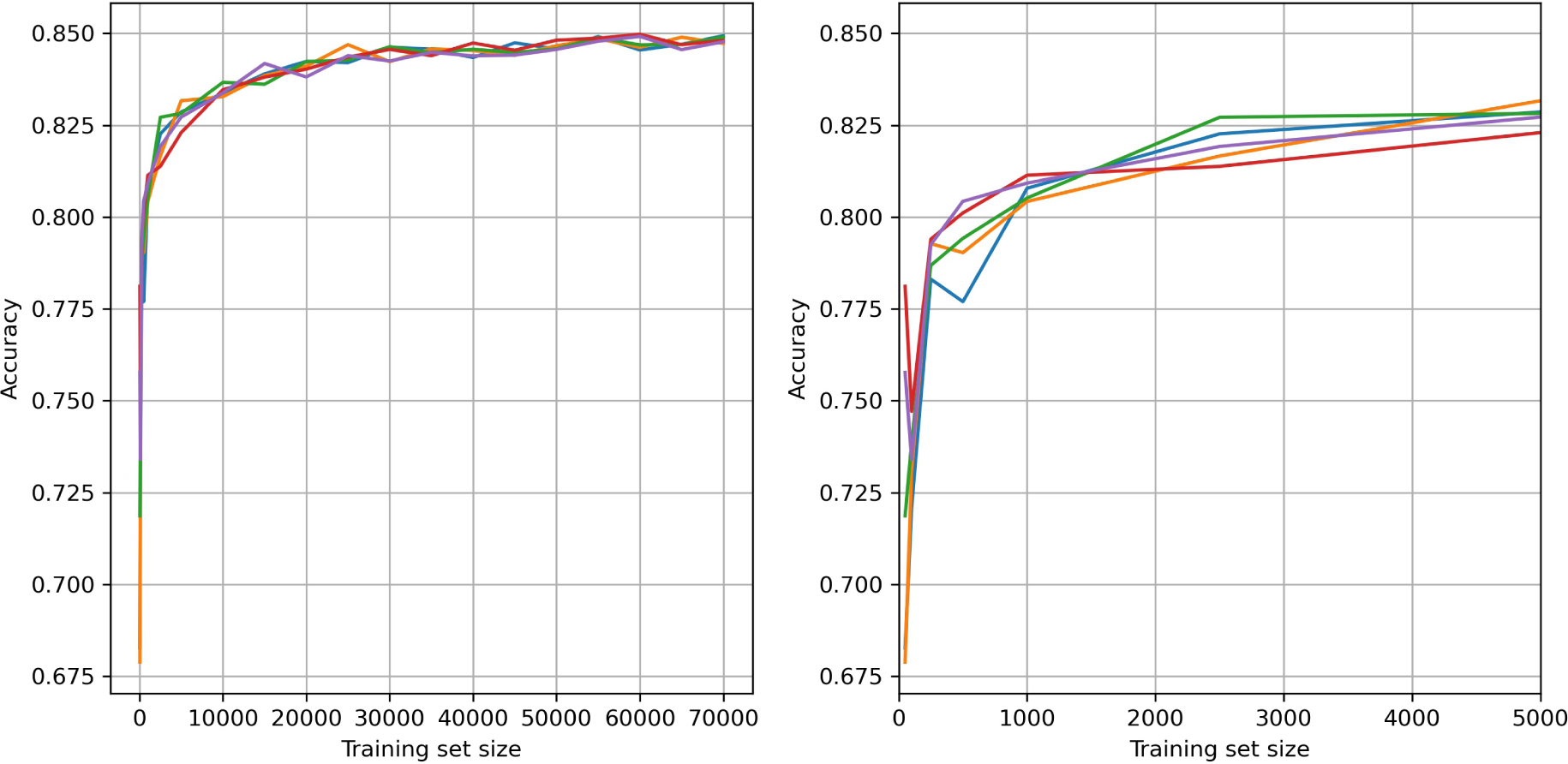
Learning curves showing the relationship between training set size and model accuracy. Left: training set size up to 70k. Right: training set size up to 5k (same results as the results on the left). Results are shown for all 5 k-fold replicates.

#### A.11 Fine-tuning of model regularisation

As hospital ID is encoded as one-hot, and there are 132 hospitals, it is possible that the effect of hospitals ID becomes ‘regularised out’, especially as for each one-hot encoded column about 99

As we are concerned with differences between hospitals, we did not want to over-regularise the model. To optimise *learning rate* we looked at the between-hospital variation of predicted thrombolysis use in a 10k cohort of patients (with the model predicting the use of thrombolysis in each hospital with the same 10k cohort). The model was trained on the remaining 78,928 patients, with varying learning rates (figure A.14 and table A.7).

**Figure A.14:**
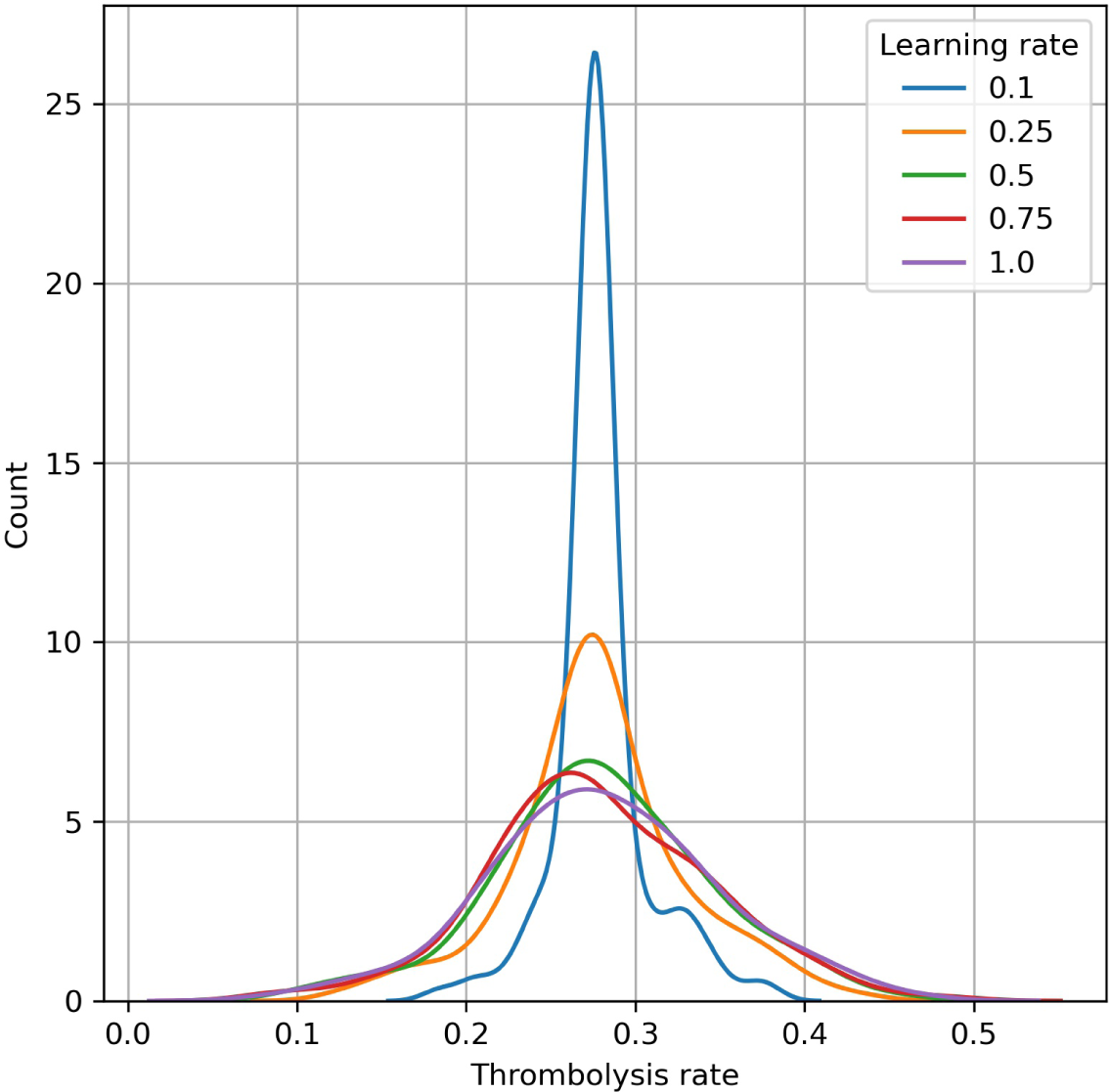
Effect of adjusting XGBoost learning rate on the distribution predicted thrombolysis use across 132 hospitals. A narrower distribution indicates that hospital thrombolysis rates are tending towards the mean thrombolysis hospital rate.

Reducing the learning rate below 0.5 led to reduced between-hospital variation in the predicted use of thrombolysis, suggesting that the effect of hospital ID was being reduced by over-regularisation.

A learning rate of 0.5 was chosen for all modelling (including the accuracy measurements above).

**Table A.7:** Statistics on the variation in predicted thrombolysis use between hospitals, with varying learning rate

| Learning rate | 0.1 | 0.25 | 0.5 | 0.75 | 1.0 |
| --- | --- | --- | --- | --- | --- |
| Mean | 0.28 | 0.28 | 0.28 | 0.28 | 0.28 |
| StdDev | 0.03 | 0.05 | 0.06 | 0.07 | 0.07 |
| Min | 0.18 | 0.13 | 0.10 | 0.09 | 0.09 |
| Max | 0.38 | 0.43 | 0.45 | 0.48 | 0.46 |

## Footnotes

a https://www.strokeaudit.org/

b https://digital.nhs.uk/data-and-information/data-tools-and-services/data-services/hospital-episode-statistics

c http://www.hra-decisiontools.org.uk/research/

d https://www.hqip.org.uk/

e https://www.strokeaudit.org/

f https://www.hqip.org.uk/

g https://digital.nhs.uk/data-and-information/data-tools-and-services/data-services/hospital-episode-statistics

h http://www.hra-decisiontools.org.uk/research/

